# A Methodological Review of Patient Healthcare-Seeking Journeys from Symptom Onset to Receipt of Care

**DOI:** 10.1101/2024.08.01.24311159

**Authors:** Charity Oga-Omenka, Angelina Sassi, Nathaly Aguilera Vasquez, Namrata Rana, Mohammad Yasir Essar, Darryl Ku, Hanna Diploma, Lavanya Huria, Kiran Saqib, Rishav Das, Guy Stallworthy, Madhukar Pai

## Abstract

**Background:** For many diseases, early diagnosis and treatment are more cost-effective, reduce community spread of infectious diseases, and result in better patient outcomes. However, healthcare-seeking and diagnoses for several diseases are unnecessarily delayed. For example, in 2022, 3 million and 5.6 million people living with TB and HIV respectively were undiagnosed. Many patients never access appropriate testing, remain undiagnosed after testing or drop out shortly after treatment initiation. This underscores challenges in accessing healthcare for many individuals. Understanding healthcare-seeking obstacles can expose bottlenecks in healthcare delivery and promote equity of access. We aimed to synthesize methodologies used to portray healthcare-seeking trajectories and provide a conceptual framework for patient journey analyses.

**Design/Methods:** We conducted a literature search using keywords related to “patient/care healthcare-seeking/journey/pathway analysis” AND “TB” OR “infectious/pulmonary diseases” in PubMED, CINAHL, Web of Science and Global Health (OVID). From a preliminary scoping search and expert consultation, we developed a conceptual framework and honed the key data points necessary to understand patients’ healthcare-seeking journeys, which then served as our inclusion criteria for the subsequent expanded review. Retained papers included at least three of these data points.

**Results:** Our conceptual framework included 5 data points and 7 related indicators that contribute to understanding patients’ experiences during healthcare-seeking. We retained 66 studies that met our eligibility criteria. Most studies (56.3%) were in Central and Southeast Asia, explored TB healthcare-seeking experiences (76.6%), were quantitative (67.2%), used in-depth, semi-structured, or structured questionnaires for data collection (73.4%). Healthcare-seeking journeys were explored, measured and portrayed in different ways, with no consistency in included information.

**Conclusions:** We synthesized various methodologies in exploring patient healthcare-seeking journeys and found crucial data points necessary to understand challenges patients encounter when interacting with health systems. and offer insights to researchers and healthcare practitioners. Our framework proposes a standardized approach to patient journey research.

**Key Questions:** *What is already known about this subject?:*

- Accessing healthcare is challenging for half of the world’s population.
- Understanding healthcare-seeking obstacles can help to expose bottlenecks in healthcare delivery and improve access.

*What does this study add?:*

- We synthesized the different methodologies used by researchers to portray healthcare-seeking trajectories.
- We also provide a conceptual framework and recommendations for patient journey analyses.

*How do the new findings imply?:*

- Our analysis revealed a lack of consistency in how patient journeys to care are represented and a notable complexity in generating insightful depictions of journeys to care.
- The use of our conceptual framework, namely the data points and indicators, could increase the reliability and generalisability patient journey analyses.

## Introduction

In 2021, 4.5 billion people or more than half of the world’s population lacked access to the healthcare they needed.^1^ Diagnoses for several diseases are unnecessarily delayed, contributing to prolonged suffering and avoidable deaths. For example, in 2022, an estimated 3 million and 5.6 million people living with TB and HIV respectively were undiagnosed. ^2,3^ A significant number of individuals with life-threatening illnesses never access appropriate testing, remain undiagnosed after testing or drop out before or shortly after treatment initiation. ^4–7^ Some reasons for these gaps in testing, diagnosis, and treatment include individual and interpersonal dynamics like lack of information about the disease and available resources, stigma, financial and cultural factors, symptom minimization, self-medication and mistrust of public sector healthcare^8–13^; as well as health system factors like poor coverage of services, low index of suspicion among providers, lengthy procedures, misdiagnosis, and poor referral mechanisms between the public and private sectors.^10,14–16^ These numbers underscore the substantial public health challenge that accessing healthcare poses for many individuals. Despite these barriers, early diagnosis and treatment remain critical for significantly improving clinical outcomes and reducing costs for both patients and the healthcare system, especially for infectious diseases like TB and HIV where early intervention is the most practical method to interrupt transmission.^10,17^

The World Health Organization (WHO) adopted the integrated people-centered health services framework in 2016 to prioritize people’s needs and expectations by engaging communities, reorienting models of care, coordinating services across sectors and strengthening governance and accountability.^18–20^ Person-centered care emphasizes treating patients as individuals and focuses on providing integrated care, patient information and support, and responding to patients’ values and preferences.^21,22^ To improve person-centered care, it is important to understand current patient journeys to care^23^. These studies shed light on the obstacles patients encounter during healthcare-seeking, expose healthcare delivery bottlenecks and promote equitable access.^24–26^ Understanding patient journeys is critical for making services more patient focused, by identifying gaps in service delivery to ensure quality care is accessible early in the healthcare-seeking process.^12,27–29^ While patient pathways analyses are emerging, lack of consistency in methods, or shared terms and frameworks makes it difficult to interpret across settings. Our review aimed to synthesize methodologies used to portray patients’ healthcare-seeking trajectories and provide a clear conceptual framework for patient journey analyses. In this review, we have explored methodologies used to highlight patient healthcare-seeking experiences from various disease contexts, including TB.

While also commonly referred to as patient pathways in literature, the healthcare-seeking journey analysis emphasized in this paper differs from the process analytics used to map and improve integration for in-hospital processes between the different departments^30–33^ or the pathway analysis to determine alignment between population health needs and available services.^34,35^ Patient journey analyses characterize and quantify the pathways to care of specific patients, detailing not only the total delays between key milestones^36–39^ but also the number and sequence of visits to healthcare providers. ^27–29,40^

Current literature has yet to explore the advantages and limitations of different methods of depicting healthcare-seeking trajectories and identifying the key variables that highlight these obstacles. The combination of conceptual and methodological reviews has been used widely in literature to critically appraise different research methods.^41–43^ A conceptual review is focused on key data points or variables and their relationships, with the goal of categorizing and describing them relevant to a particular topic.^44–46^ Methodological reviews differ from a traditional systematic review by prioritizing *methods* over *results*,^47–50^ synthesizing study quality by examining design, data collection and analysis.^47,48^ This review aims to synthesize methods for portraying patients’ trajectories for TB and other diseases and propose a conceptual framework for analyzing these journeys.

## Methods

### Review Objectives

The primary aim of our review was to assess relevant current practices, methodologies and relevant data points used to map patient care trajectories for TB and related diseases, focusing on the patient healthcare-seeking journey from symptom onset to care outcome across diverse healthcare settings and conditions. The research question was: ‘How do the methodologies employed for patient journey analysis (PJA) differ, what are their strengths and limitations?’.

### Review Approach

We use the conceptual and methodological review approaches, which adapt their scope and methods during the process.^48,50^ We used an iterative approach, using literature review and expert consultations to understand the patient healthcare-seeking journey. This entailed extensive database searches and consultations with experts in tuberculosis care and social science, and refining review findings based on iterative feedback.

### Search Strategy and Framework Development

Our process comprised of three steps: (1) A scoping literature search to identify relevant data points and methods; (2) Expert consultation to identify conceptual papers and recommended measures, followed by the development of conceptual map; and (3) An updated literature search targeting papers that included identified data points.

#### Initial scoping literature search

We performed a scoping search on PubMed, Web of Science, Scopus, and Google Scholar for publications on healthcare-seeking trajectories and delays related to TB diagnosis or treatment. The initial search was run in November 2021 (updated later, see below) and was restricted to peer-reviewed publications in the English language with no date restrictions. We also consulted with TB care experts to identify additional papers.

#### Development of the conceptual framework

Our conceptual framework was derived from the initial literature search followed by expert consultations. The lead author reviewed the literature and identified multiple criteria— variables, factors, and themes—that describe and influence patients’ journeys to care. The research team had several meetings to discuss, clarify, and define the key features to be included in the framework, culminating in developing the first draft. To validate the framework, the team sought expert consultation from experienced researchers and healthcare professionals in the field of TB and HIV care in several meetings, workshops, and conferences to identify additional data points, and ensure a comprehensive understanding of key methodological considerations. The conceptual framework, identifying key variables or themes for two care outcomes – diagnosis and treatment, was refined based on feedback from these interactions and used to define the inclusion criteria. As a result of this process, we decided to expand our initial literature search to cover methodologies used to describe patient healthcare-seeking journeys for infectious or pulmonary diseases, or diseases with non- specific symptoms or lengthy diagnostic procedures – including HIV, COVID, malaria and cancer, to strengthen the insights gained from this methodological review beyond what could be inferred from TB research alone. Defining the conceptual framework helped with the rest of the methodological review to identify the lack of consistency in methods and analytic approaches.

#### Updated literature search and inclusion criteria

We systematically searched PubMed, CINAHL, Web of Science, Global Health (OVID), and Scopus for publications on healthcare-seeking trajectories, delays to diagnosis or treatment, and access to diagnosis or treatment, for TB, HIV, malaria, COVID-19, and other respiratory and lung disorders. The initial search of November 2021 was updated in April 2022 and searched again in May 2023. Our search strategy can be found in *Supplement 1*.

To be included in our review, studies had to include at least three of the identified data points for assessing patient journeys within the healthcare system in low- and middle-income countries (*Table 1*). We excluded case studies of individuals; non-scientific publications such as opinions or editorials; systematic reviews and meta-analyses; studies on extrapulmonary tuberculosis (EPTB), latent TB, and pediatric TB (children under 15 years); studies focusing solely on the cascade of care; articles written in languages other than English or French; and articles that did not provide a full report of the results of an experimental study (abstract, reviews, commentaries, proposals, methodology papers, or case study).

**Table 1:** The key data points of patient healthcare-seeking journey.

| Reference points | Data points | Purpose | How to measure (quantitatively and qualitatively) | Studies |
| --- | --- | --- | --- | --- |
| Symptoms recognition | Date symptoms started | To determine the time when symptoms started or when patients recognized their symptoms | This is a date variable. Can also be expressed in time increments (e.g., weeks, months, etc.). | 28,59-91 |
|  | Type of provider first visited after symptom onset | To identify the first healthcare provider that patients sought for their symptoms until diagnosis after recognition | E.g. physician, nurse, pharmacist, laboratory technician, medicine vendor | 28,60-65,68,69,79,82,84-88,90-106 |
|  | Sector of facility first visited | To specify type of facility for that encounter | E.g. private sector, public sector, or informal sector | 28,60-65,68,79,84-88,90-106 |
| Symptom recognition to Diagnosis | Date of (or time to) diagnosis | To determine how long it took for patients to get diagnosed | This is a date variable. Can also be expressed in time increments (e.g., weeks, months, etc.). | 28,59-91,93,96 |
|  | Number of provider encounters to diagnosis | To identify the number of visits it took for patients to go from the first encounter until diagnosis | 1st encounter, 2nd encounter, etc. | 28,40,59-64,67,69,71-73,77,79,81,83,84,86,88,90-93,96-98,102,103,105,107-110 |
|  | Type of provider for each encounter to diagnosis | To specify the type of provider the patients saw at each encounter | E.g. physician, nurse, pharmacist, laboratory technician, medicine vendor | 28,60,63-65,68,69,79,82,84-88,90-106,111 |
|  | Sector of facility for each encounter | To specify type of facility for where patients were diagnosed | E.g. private sector, public sector, or informal sector | 28,60,63-65,68,79,84-88,90-106,111 |
|  | Proportions of patients per number of encounters to diagnosis | To determine the proportion of patients who had that many encounters. Also helps to identify missed opportunities for that provider | Expressed as % of 1st encounter, 2nd encounter, etc.; or number of patients at each reference point, e.g. "20% of patients had 1-2 encounters, 30% had 3 encounters..." | 28,60,62,63,66,73,77,79,83,87,88,91,92,95,98,102,103,108,110,112 |
| Diagnosis to treatment | Date of (or time to) treatment initiation | To determine how long it took for patients to start treatment once diagnosed | This is a date variable. Can also be expressed in time increments (e.g., weeks, months, etc.). | 28,60-66,68-73,75-77,80,81,85-87,89,93-96,103,105,112-118 |
|  | Number of provider encounters to treatment initiation | To identify the number of visits to a provider that it took for patients to start treatment after diagnosis. | 1st encounter, 2nd encounter, etc. | 60,63,72,75-77,81,85,87,93,94,96,99,101,103,105,110,114,115,117 |
|  | Type of provider for each encounter to treatment initiation | To specify the type of provider the patients saw at each encounter | E.g. physician, nurse, pharmacist, laboratory technician, medicine vendor | 60-63,85,87,88,90,91,93,94,96,99,101,103,105,111,113,119-121 |
|  | Sector of facility for each encounter to treatment initiation | To specify type of facility where patients were started on treatment | E.g. private sector, public sector, or informal sector | 60,63,85,87,88,90,91,93,94,96,99,101,103,105,119-121 |
|  | Proportions of patients per number of encounters to treatment initiation | To determine the proportion of patients who had that many encounters. Also helps to identify missed opportunities for that provider | Expressed as % of 1st encounter, 2nd encounter, etc.; or number of patients at each reference point, e.g. "20% of patients had 1-2 encounters, 30% had 3 encounters..." | 62,63,76,83,85,87,93,95,105,110,112,120 |

References retrieved from all databases were imported into Covidence. Title and abstract screening were carried out by five members of the team (NAV, COO, DK, AS, and MYE) and full-text screening by four members of the team (COO, LH, NAV, and MYE), meeting regularly to review progress and resolve conflicts. Agreement on inclusion between two reviewers was required for inclusion into the study. Conflicts were resolved by a third reviewer when necessary.

There are no clear guidelines for appraising the quality and risk of bias for methodological reviews.^47,48^ The focus of the appraisal process for this type of review should be on distinguishing between papers stemming from flawed empirical studies and those presenting well-argued theories.^47^ We used the JBI Critical Appraisal Checklists^51^ for each applicable study type (qualitative, cross-sectional, cohort, or prevalence study) and the Mixed Methods Appraisal Tool^52^ for mixed-methods studies to evaluate the methodological rigor and reliability of the included studies and identify potential biases and limitations within the studies. Quality assessment was carried out by six members of the team (AS, NAV, RD, DK, MYE, and KS) using an electronic form which was piloted by AS and NR (*Supplement 3)*. Quality assessment for each paper was done by two independent reviewers and answers were finalized during a consensus meeting led by AS and MYE.

## Data Availability

All data produced in the present work are contained in the manuscript

## Data extraction

Data extraction was done by six members of the team (AS, MYE, NR, DK, HD, and CO) using an electronic data extraction form hosted on Covidence (*Supplement 2*). The data extraction form was piloted by CO, NAV, LH, and NR prior to data extraction. The data extraction form aimed to collect general information (author name, publication year, country/setting, disease type, study design/data sources, study type and sample size), the journey criteria included for diagnosis and treatment, a general description of the methodology and analysis for each study, as well as strengths and limitations. Data extraction for each paper was done by two independent reviewers, and answers were finalized during a consensus meeting led by AS and MYE.

## Ethical considerations

No human participants were involved in this review.

## Results

### Definition of Key Data points and the Conceptual Map

The care continuum describes the sequence of activities needed to be fully engaged in clinical care for diseases like TB and HIV that requiring lengthy health system procedures to achieve an outcome.^53–55^ Our patient journey framework, developed after the initial review of literature, highlights the patient healthcare-seeking trajectory across three reference points: symptom recognition, diagnosis, and treatment initiation. Our review illustrates how patients consult various types of providers in differing numbers as they seek care between these reference points. To characterize each visit within a patient’s journey, we identified five key data points: (1) the date of the first visit relative to symptom onset, (2) the type of provider or facility visited (e.g., chemist, general practitioner, specialist, laboratory, hospital), (3) the sector of the provider or facility (for-profit, non-profit, public), (4) the sequence of the visit (first visit, second visit, etc.), and (5) the outcome of the visit (no outcome, diagnosis, treatment initiation). Using these data points, we can compute various indicators for both individual patients and aggregated patient populations. These measures include (1) the number of days and (2) visits until care outcome (diagnosis or treatment initiation), (3) the proportion of visits to different types of providers, (4) the contribution of various provider types to care outcome (diagnosis or treatment) as well as to (6) delays or missed opportunities, and (7) the number of patients exhibiting different patterns or sequences of provider visits. This comprehensive analysis can inform strategies to optimize patient care experiences and reduce delays in diagnosis and treatment.

Our conceptual framework (*Figure 1*) illustrates the steps in a patient journey and the time delays between key milestones of symptom recognition, diagnosis, and treatment. Starting with symptom recognition, factors like symptom minimization and lack of knowledge of services have been shown to influence healthcare-seeking delays, while the type and sector of first provider have been shown to influence number of subsequent encounters before and timeliness of diagnosis.^10,11,56,57^ Several health system factors have been shown to influence the timing and number of encounters between the reference points of diagnosis and treatment.^10,11,58^ Delays are conceptualized as healthcare-seeking, provider and treatment delays, with diagnostic delays being the sum of healthcare-seeking and provider delays, and health system delays representing the sum of provider and treatment delays.^27,29^

**Figure 1:**
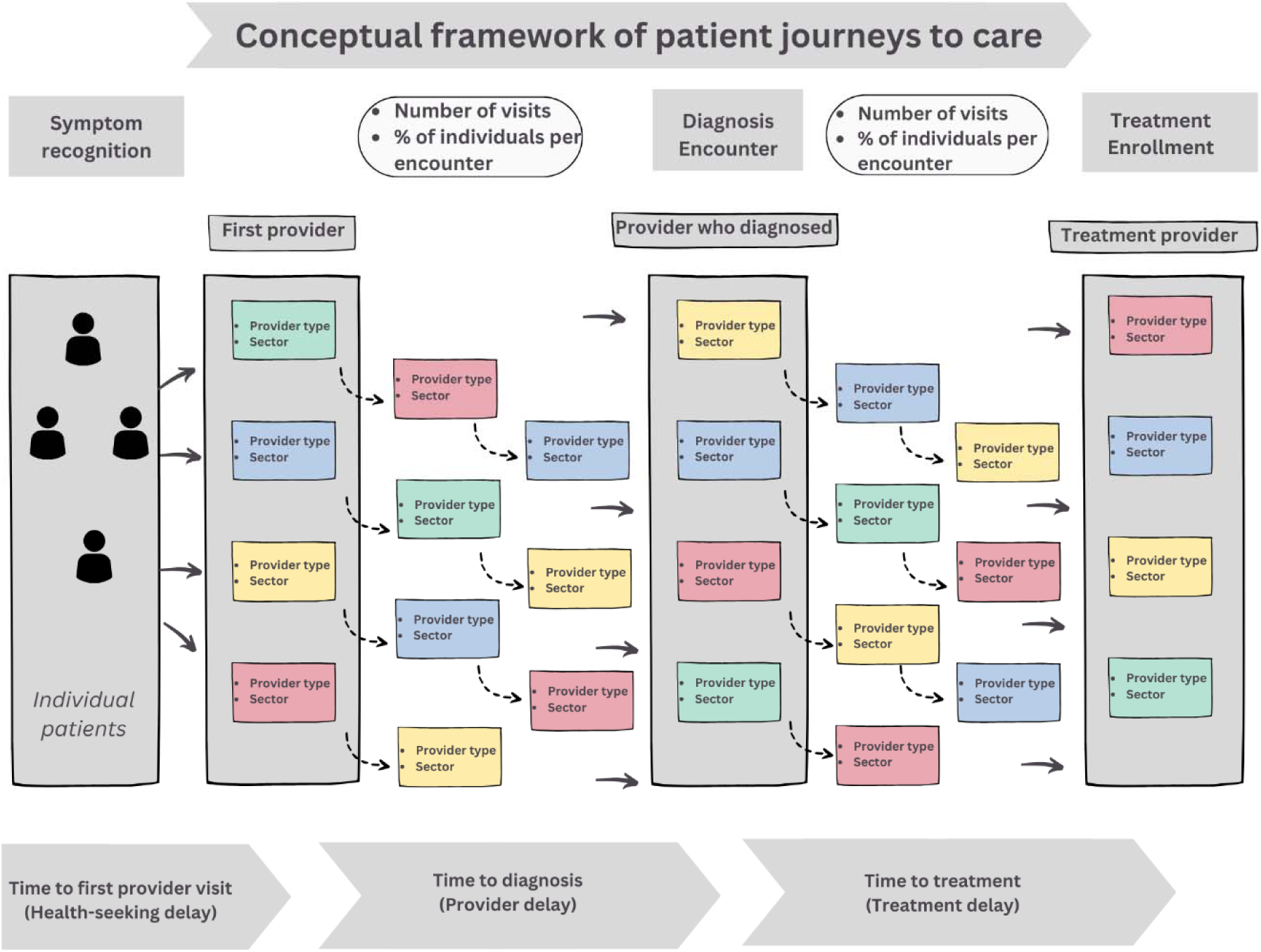
Conceptual framework of healthcare-seeking journeys.

### Characteristics of Included Studies

The PRISMA flow diagram (*Figure 2*) shows search results from the five main databases. Out of the 12160 studies whose titles and abstracts were initially screened from those databases, 297 studies were assessed for retrieval and eligibility, 231 were excluded through full-text review, and 66 studies were retained that met our eligibility criteria. Each of these 66 studies (our unit of analysis) was a patient journey analysis publication.

**Figure 2:**
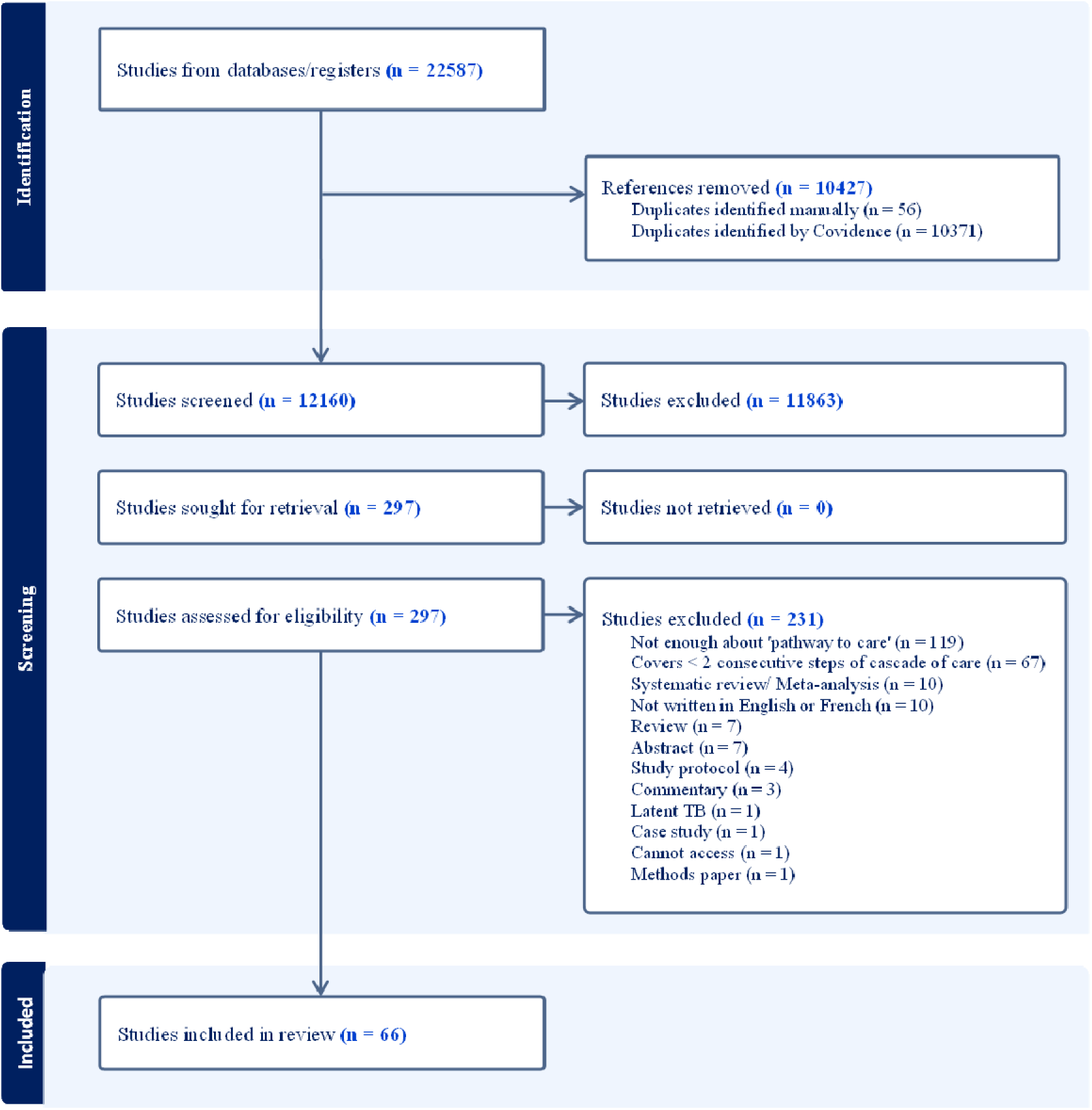
Prisma diagram of study selection process.

Out of the 66 retained studies, we assessed 21 to be of high quality, 37 to be medium quality, and 8 to be of low quality. There were no major differences in the methods for the studies assessed with different qualities.

*Table 2* summarizes journey methodology characteristics of included studies. Many studies were quantitative (82%), were conducted in lower middle-income countries (56%), in Asia (59%), and covered TB care journeys (67%). Other diseases include the following: cancer (6.1%), acute febrile illnesses (4.5%), malaria (4.5%), chest symptomatic (3.0%), multiple infectious diseases (3.0%), non-communicable diseases (3.0%), sexually transmitted infections (3.0%), under-five mortality (3.0%), and COVID-19 (1.5%).

**Table 2:**
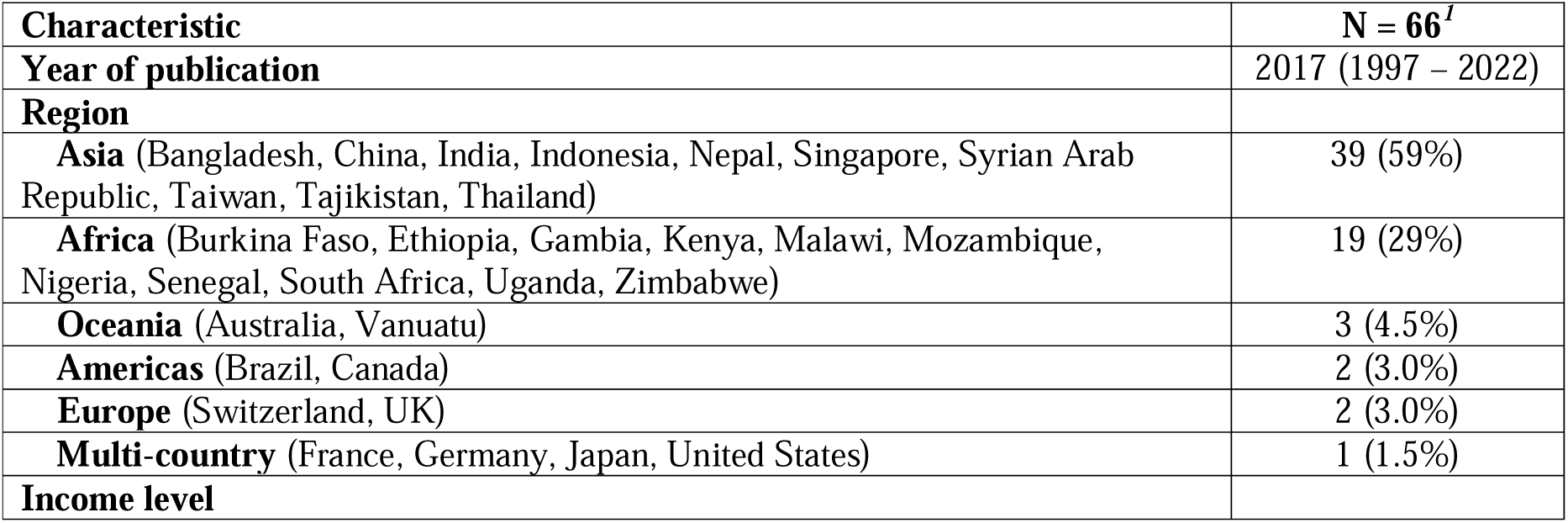
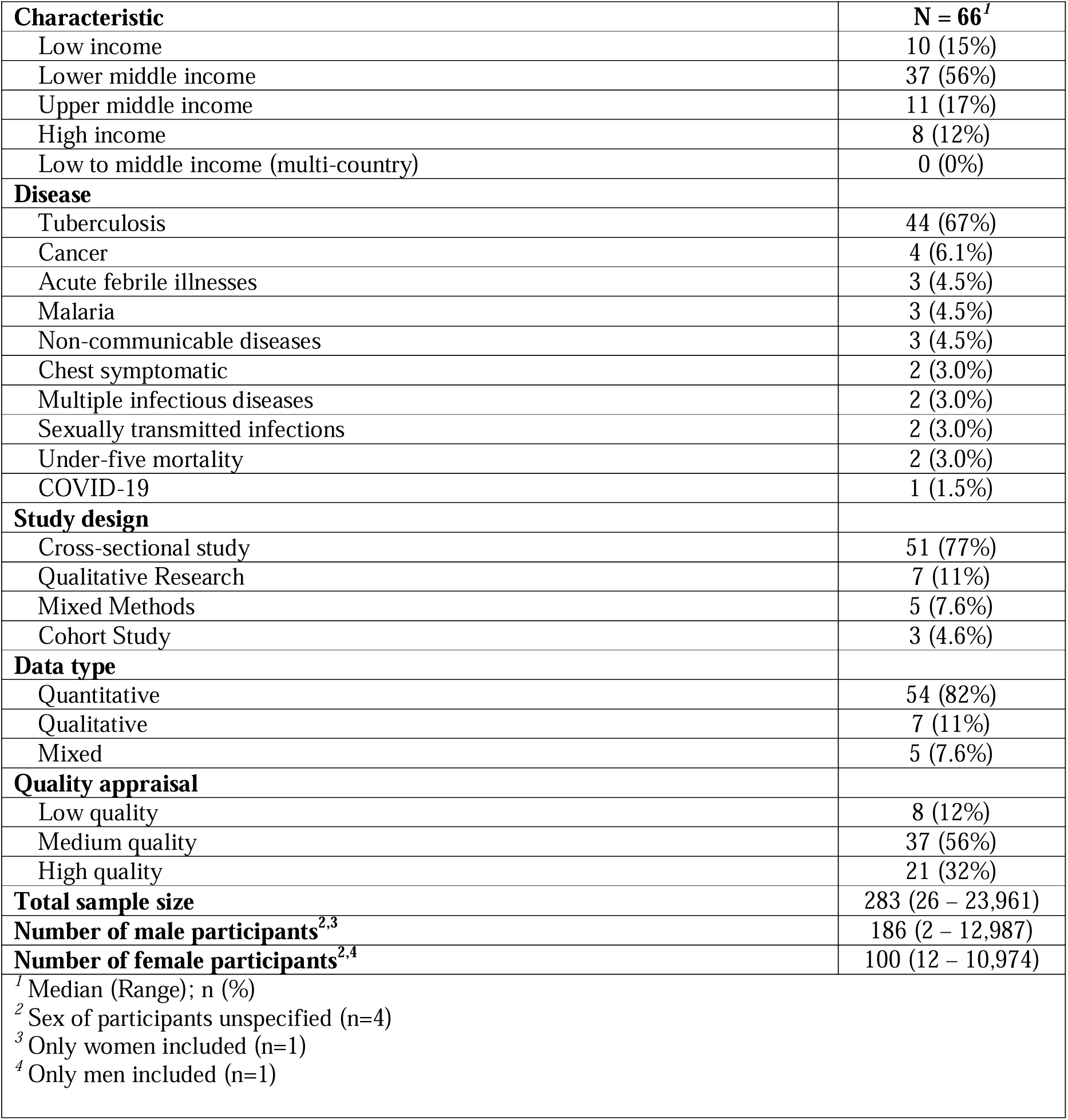
Characteristics of included manuscripts.

*Figure 3a* summarizes the distribution of patient healthcare-seeking data points aggregated across all included studies. The majority included information on location/provider where treatment was initiated (67%), sector of the providers visited (61%), time to diagnosis (56%), location/provider where a diagnosis was made (55%), number of provider encounters to diagnosis (53%), and type of the providers visited (50%). Proportions of patients per number of encounters to diagnosis (33%) and treatment (20%), and number of provider encounters to treatment (30%) were less commonly explored in the included papers. *Figure 3b* shows that most of the patient healthcare-seeking data points were reported within studies the four disease groups. Type of facility/provider where treatment was started is more frequently found in articles focused on malaria and fevers and other infectious diseases compared to other disease types. Most categories of data were available in all types of study design (*Figure 3b*).

**Figure 3:**
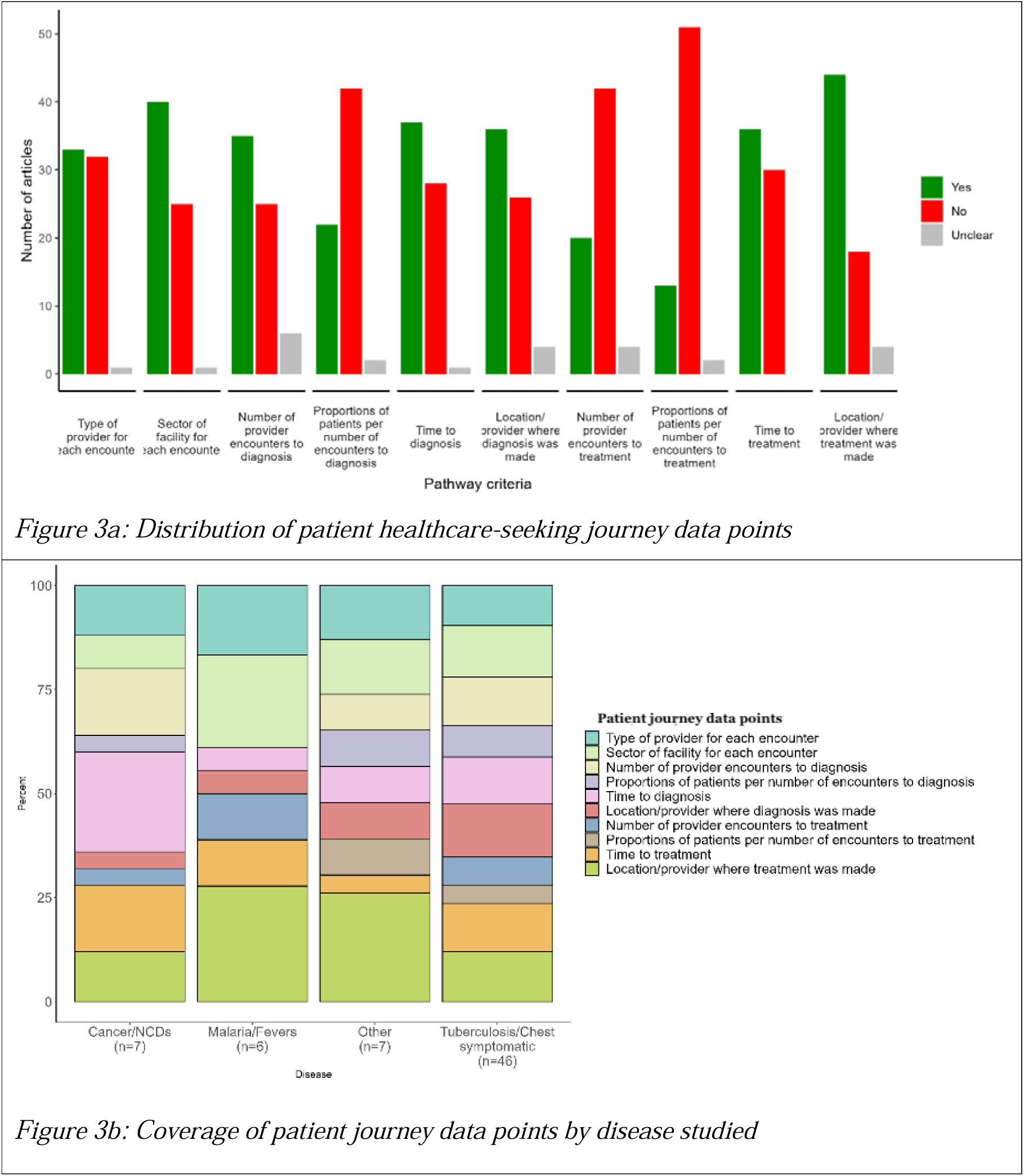
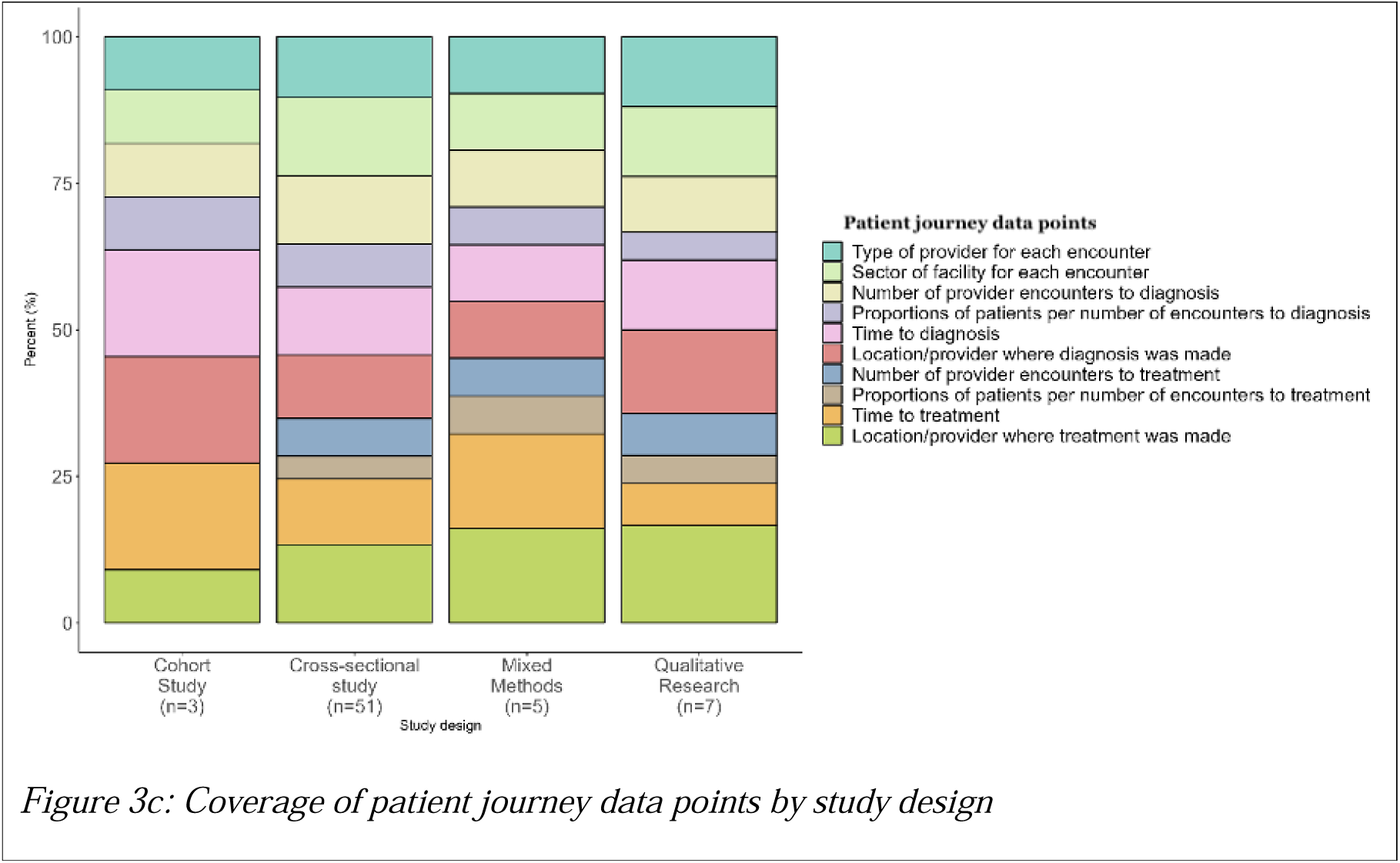
Distribution of journey data points across all included studies, by disease types and study designs.

*Table 3* details the depictions of healthcare-seeking journeys and statistical methods used in the included studies. The most common forms of depicting patient healthcare-seeking journeys were tables quantifying delays and/or journeys (48%) and flowcharts (48%). Descriptive statistics (88%), multivariate regressions (45%), chi-squared test (45%), and logistic regression (36%) were the most frequently used statistical methods.

**Table 3:** Journey depictions and statistical methodologies.

| Characteristic | N = 66<sup>†</sup> |
| --- | --- |
| **Depictions of patient journeys to care** |  |
| Tables quantifying delays/journeys | 32 (48%) |
| Flow chart | 32 (48%) |
| Bar chart | 12 (18%) |
| Authors' own depiction (other) | 9 (14%) |
| Matrix representing patient journeys | 6 (9.1%) |
| Diagram summarizing types of delays | 5 (7.6%) |
| Sankey chart | 3 (4.5%) |
| Scatter plot | 1 (1.5%) |
| **Statistical methodologies used** |  |
| **Main statistical method used per article** |  |
| Logistic regression only | 18 (27%) |
| Chi-square test only | 7 (11%) |
| Independent T-test and Chi-square test only | 7 (11%) |
| More than one regression method | 7 (11%) |
| Thematic analysis only | 7 (11%) |
| Descriptive statistics only | 5 (7.6%) |
| Independent T-test only | 5 (7.6%) |
| Mixed methods | 4 (6.1%) |
| Linear regression only | 3 (4.5%) |
| Analysis of variance (ANOVA) test only | 1 (1.5%) |
| Quantile regression only | 1 (1.5%) |
| Survival analysis only | 1 (1.5%) |
| Descriptive statistics | 58 (88%) |
| Chi-squared test | 30 (45%) |
| Independent T-test | 22 (33%) |
| Fisher's exact test | 8 (12%) |
| Mann-Whitney test | 8 (12%) |
| Kruskal-Wallis test | 4 (6.1%) |
| Analysis of variance (ANOVA) | 3 (4.5%) |
| Multivariate regression (all types) | 30 (45%) |
| Logistic regression | 24 (36%) |
| Linear regression | 8 (12%) |
| Survival analysis | 3 (4.5%) |
| Poisson regression | 2 (3.0%) |
| Quantile (median) regression | 2 (3.0%) |
| Thematic analysis of qualitative data | 11 (17%) |
| <sup>l</sup> n (%) |  |

Table 4 depicts the outcome measures and independent variables used in the included articles. Overall, patient delay (58%) was the most common outcome measure for the 66 included studies. Over one-third (42%) of included articles did not report all possible delays that could have been reported in the manuscript; for example, if the authors reported diagnostic delay (defined^27,29^ as the number of days between the onset of symptoms and the date of diagnosis and composed of patient delay and provider delay) but did not also report both patient and provider delay. Other than overall delay, the most reported outcome measures of patient journeys were number of visits to diagnosis (36%), sector of facility and type of provider for all instances of healthcare-seeking (36% and 33%, respectively), and type of provider for initial healthcare-seeking visit (32%). Time to initial healthcare-seeking was reported in 29% of included studies. Patient age (97%) and sex or gender (92%) were the two most used independent variables among the 66 included studies, followed by level of education (62%) and occupation (59%). Among patient health status and patient behavior variables, type of disease (30%), presence of any comorbidities (27%), and HIV status (27%) were often included as independent variables. Some patient healthcare-seeking journey characteristics were used as independent variables in analyses of delays, for example, number of visits (30%), sector of facility (39%) and type of provider (38%) of initial healthcare-seeking visit.

**Table 4:**
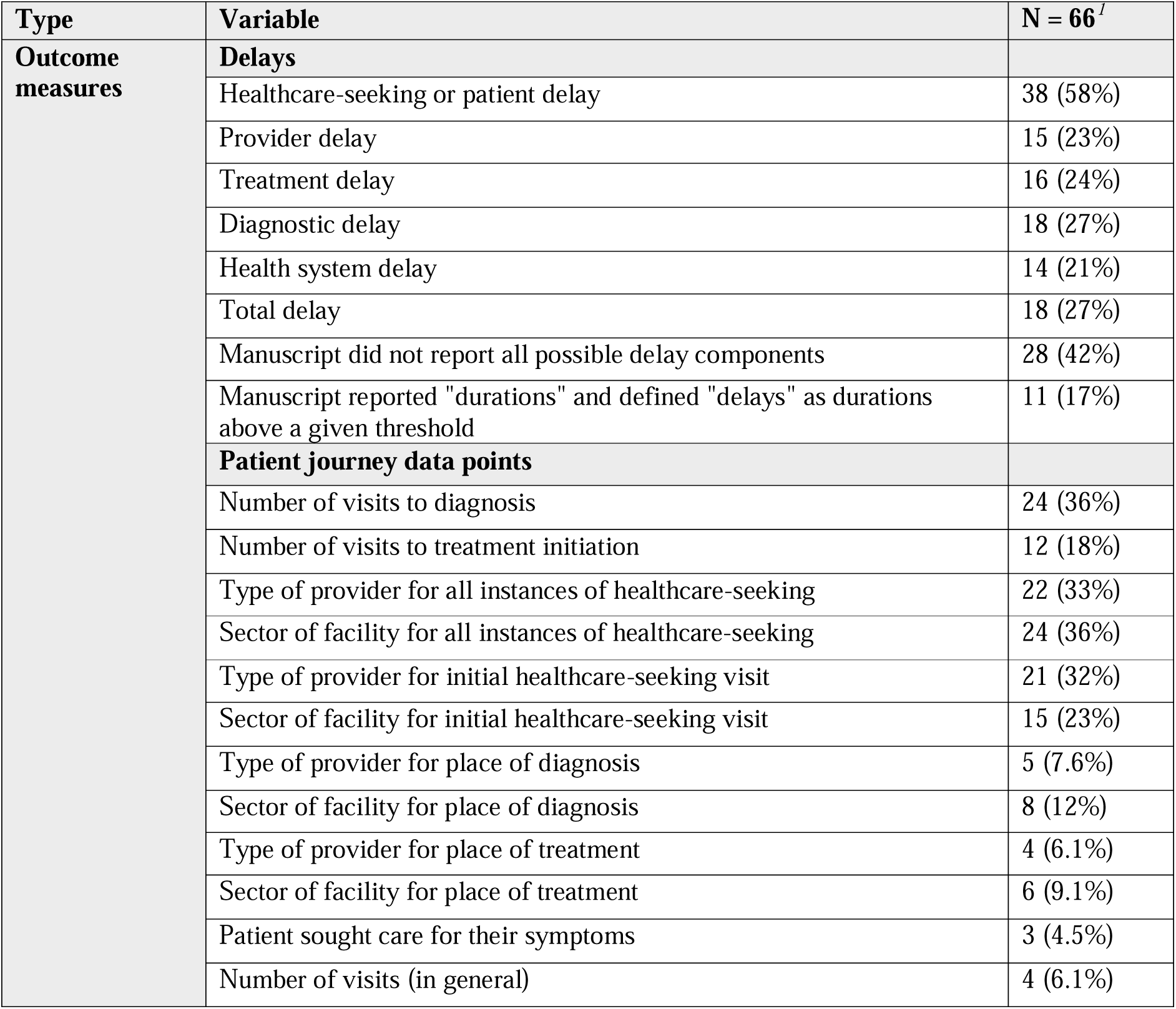
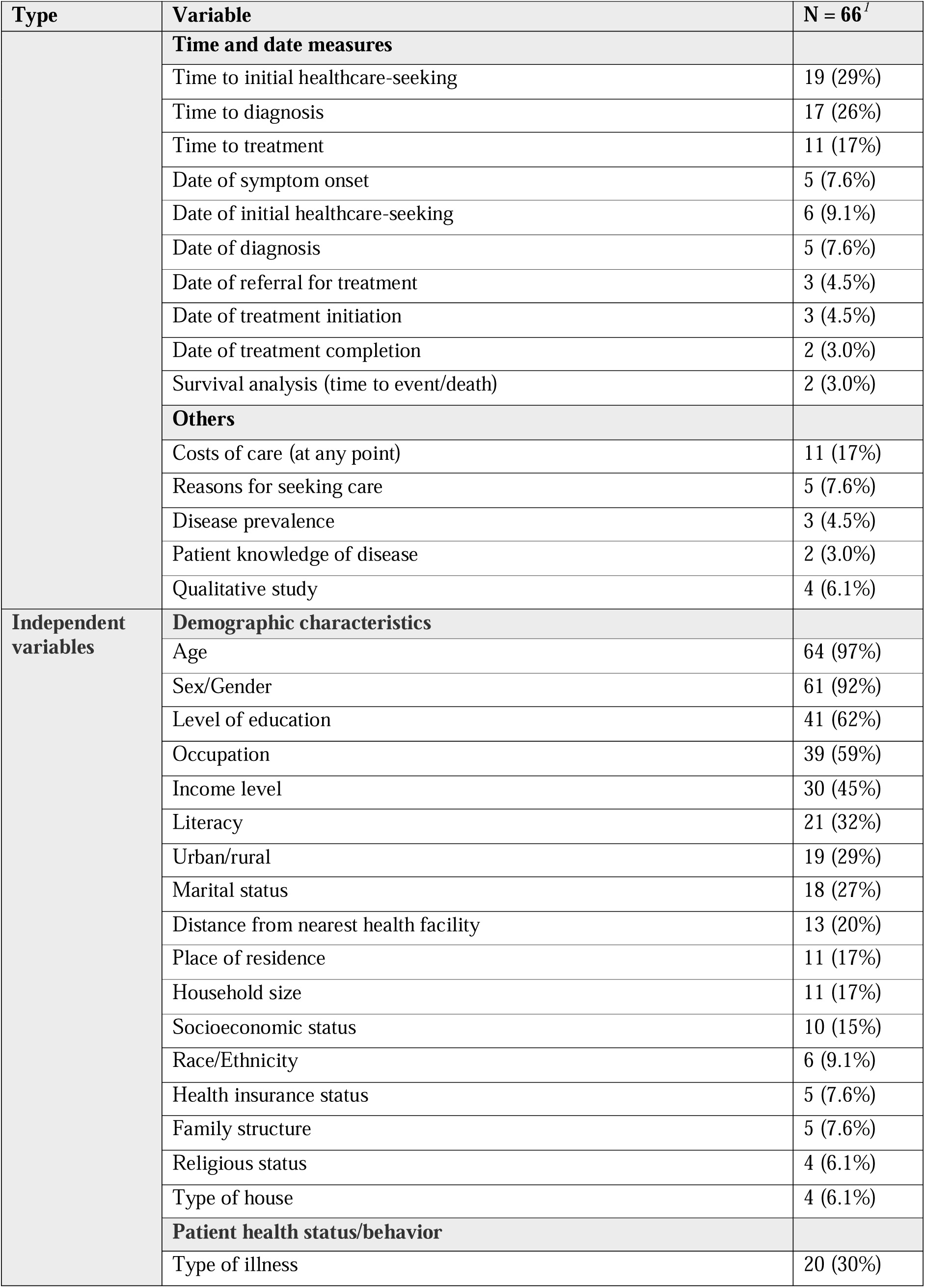
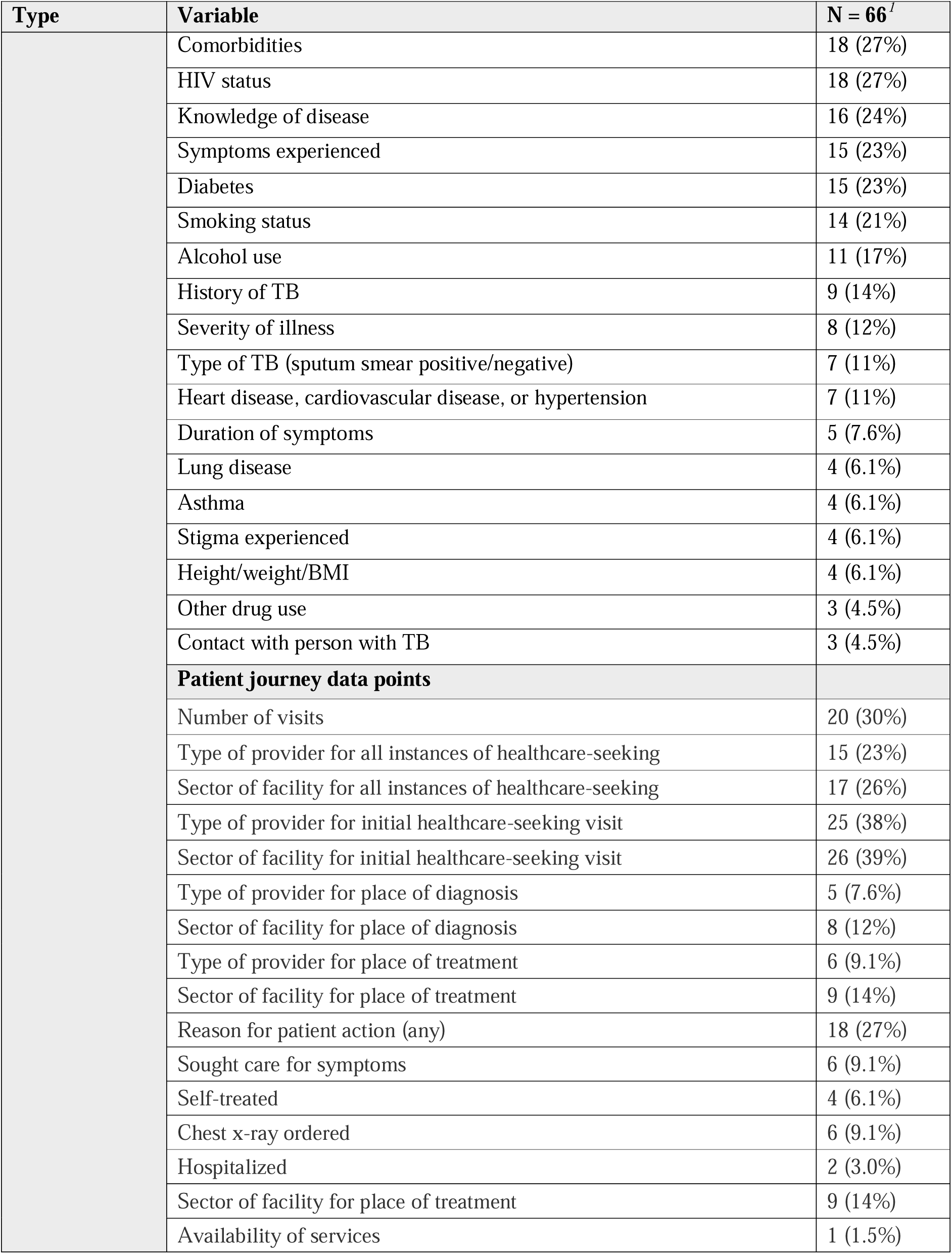
Outcome measures and independent variables used.

| Type | Variable | N = 66<sup>l</sup> |
| --- | --- | --- |
| Outcome measures | **Delays** |  |
|  | Healthcare-seeking or patient delay | 38 (58%) |
|  | Provider delay | 15 (23%) |
|  | Treatment delay | 16 (24%) |
|  | Diagnostic delay | 18 (27%) |
|  | Health system delay | 14 (21%) |
|  | Total delay | 18 (27%) |
|  | Manuscript did not report all possible delay components | 28 (42%) |
|  | Manuscript reported "durations" and defined "delays" as durations above a given threshold | 11 (17%) |
|  | **Patient journey data points** |  |
|  | Number of visits to diagnosis | 24 (36%) |
|  | Number of visits to treatment initiation | 12 (18%) |
|  | Type of provider for all instances of healthcare-seeking | 22 (33%) |
|  | Sector of facility for all instances of healthcare-seeking | 24 (36%) |
|  | Type of provider for initial healthcare-seeking visit | 21 (32%) |
|  | Sector of facility for initial healthcare-seeking visit | 15 (23%) |
|  | Type of provider for place of diagnosis | 5 (7.6%) |
|  | Sector of facility for place of diagnosis | 8 (12%) |
|  | Type of provider for place of treatment | 4 (6.1%) |
|  | Sector of facility for place of treatment | 6 (9.1%) |
|  | Patient sought care for their symptoms | 3 (4.5%) |
|  | Number of visits (in general) | 4 (6.1%) |

| Type | Variable | N = 66<sup>I</sup> |
| --- | --- | --- |
|  | **Time and date measures** |  |
|  | Time to initial healthcare-seeking | 19 (29%) |
|  | Time to diagnosis | 17 (26%) |
|  | Time to treatment | 11 (17%) |
|  | Date of symptom onset | 5 (7.6%) |
|  | Date of initial healthcare-seeking | 6 (9.1%) |
|  | Date of diagnosis | 5 (7.6%) |
|  | Date of referral for treatment | 3 (4.5%) |
|  | Date of treatment initiation | 3 (4.5%) |
|  | Date of treatment completion | 2 (3.0%) |
|  | Survival analysis (time to event/death) | 2 (3.0%) |
|  | **Others** |  |
|  | Costs of care (at any point) | 11 (17%) |
|  | Reasons for seeking care | 5 (7.6%) |
|  | Disease prevalence | 3 (4.5%) |
|  | Patient knowledge of disease | 2 (3.0%) |
|  | Qualitative study | 4 (6.1%) |
| **Independent variables** | **Demographic characteristics** |  |
|  | Age | 64 (97%) |
|  | Sex/Gender | 61 (92%) |
|  | Level of education | 41 (62%) |
|  | Occupation | 39 (59%) |
|  | Income level | 30 (45%) |
|  | Literacy | 21 (32%) |
|  | Urban/rural | 19 (29%) |
|  | Marital status | 18 (27%) |
|  | Distance from nearest health facility | 13 (20%) |
|  | Place of residence | 11 (17%) |
|  | Household size | 11 (17%) |
|  | Socioeconomic status | 10 (15%) |
|  | Race/Ethnicity | 6 (9.1%) |
|  | Health insurance status | 5 (7.6%) |
|  | Family structure | 5 (7.6%) |
|  | Religious status | 4 (6.1%) |
|  | Type of house | 4 (6.1%) |
|  | **Patient health status/behavior** |  |
|  | Type of illness | 20 (30%) |
|  | Comorbidities | 18 (27%) |
|  | HIV status | 18 (27%) |
|  | Knowledge of disease | 16 (24%) |
|  | Symptoms experienced | 15 (23%) |
|  | Diabetes | 15 (23%) |
|  | Smoking status | 14 (21%) |
|  | Alcohol use | 11 (17%) |
|  | History of TB | 9 (14%) |
|  | Severity of illness | 8 (12%) |
|  | Type of TB (sputum smear positive/negative) | 7 (11%) |
|  | Heart disease, cardiovascular disease, or hypertension | 7 (11%) |
|  | Duration of symptoms | 5 (7.6%) |
|  | Lung disease | 4 (6.1%) |
|  | Asthma | 4 (6.1%) |
|  | Stigma experienced | 4 (6.1%) |
|  | Height/weight/BMI | 4 (6.1%) |
|  | Other drug use | 3 (4.5%) |
|  | Contact with person with TB | 3 (4.5%) |
|  | **Patient journey data points** |  |
|  | Number of visits | 20 (30%) |
|  | Type of provider for all instances of healthcare-seeking | 15 (23%) |
|  | Sector of facility for all instances of healthcare-seeking | 17 (26%) |
|  | Type of provider for initial healthcare-seeking visit | 25 (38%) |
|  | Sector of facility for initial healthcare-seeking visit | 26 (39%) |
|  | Type of provider for place of diagnosis | 5 (7.6%) |
|  | Sector of facility for place of diagnosis | 8 (12%) |
|  | Type of provider for place of treatment | 6 (9.1%) |
|  | Sector of facility for place of treatment | 9 (14%) |
|  | Reason for patient action (any) | 18 (27%) |
|  | Sought care for symptoms | 6 (9.1%) |
|  | Self-treated | 4 (6.1%) |
|  | Chest x-ray ordered | 6 (9.1%) |
|  | Hospitalized | 2 (3.0%) |
|  | Sector of facility for place of treatment | 9 (14%) |
|  | Availability of services | 1 (1.5%) |

## Discussions

In our review, we aimed to understand and characterize methodologies used to assess patient journeys to care. Understanding patient journeys helps pinpoint bottlenecks and interventions to minimize missed diagnostic and treatment opportunities among various types of healthcare providers, improve hospital coordination and reduce overall delays in care. Moreover, accurately characterizing aggregate patient journeys between different locations or across different times enhances progress monitoring and policymaking. This requires developing better, consistent metrics and more insightful data visualizations.

Our analysis revealed a lack of consistency in how patient journeys to care are represented and a notable complexity in generating insightful depictions of journeys to care, indicating the challenges in using data to guide healthcare interventions effectively. The scarcity of tools and clear guidelines for analyzing and presenting such complex data further complicates this task, highlighting an urgent need for simple, open-access tools and standardized analytical methods to improve our understanding of patient healthcare-seeking journeys.

We posit that our proposed conceptual framework, identified data points and indicators address this knowledge gap effectively, offering guidance to improve consistency in the analysis of patient journey or healthcare-seeking experiences.

The reviewed papers typically addressed an average of four identified key data points, with patient delays, and the type or sector of facility where the patient was diagnosed or treated being the most often examined. We also noted that the number of provider visits needed for diagnosis or treatment were only reported in 24% and 12% respectively. In our view, the number of visits to diagnosis and treatment are among the most important data points for understanding patient care barriers, as both give us an insight into delays as well as the direct and indirect costs of care. Overall, as none of these factors alone fully characterize patient experiences, or highlight missed diagnostic and treatment opportunities, reporting all data points relevant to the outcome under review (diagnosis or treatment) is crucial for a comprehensive view of patient care journeys. Thus, our methodological review highlighted the key features of patient trajectories (*Table 1*) that offer insights into the missed opportunities. Additionally, we recommend additional statistical analyses to identify risk factors for having more provider visits or longer delays in the patient care continuum using patients’ sociodemographic characteristics and health status (*Table 4*). We propose the need for an open source analysis package for Patient Journey Analyses.

### Strengths and limitations of study designs

We assessed the strengths and limitations for each study design. Cross-sectional studies provide overview of patient proportions and care-seeking behaviours and quantify patient proportions between visits or delays. Still, varying tools and measures across studies can make collating results challenging and lead to inconsistencies. Cohort studies excel in examining care-seeking behaviors over time and identifying risk factors for delays, providing insights into temporal relationships and complex journeys, though they require large sample sizes and are also vulnerable to recall bias, especially in studies of diseases with low prevalence.

Despite being more resource-intensive, mixed methods research combines quantitative data with rich qualitative insights from key informants. Qualitative research delves deep into the experiences and obstacles within patient journeys, offering flexibility in adapting to different contexts, but is similarly limited by recall bias, high costs, and small sample sizes.

### Implications for Future Research

Patient journey analyses are prone to potential inconsistencies in data collection, particularly regarding sampling of patients, measuring encounters by number of providers or visits, and the measurement of time reference points (symptom onset, diagnosis, and treatment). Future research should focus on developing clear, practical recommendations for conducting patient journey analyses and creating open-access tools to overcome methodological and resource limitations. These advancements are crucial for researchers and teams aiming to analyze patient journeys effectively to identify and address care bottlenecks. Learning from our experiences and adopting these strategies will improve the reliability and comparability of patient journey analyses in future studies.

To mitigate recall bias, researchers need to pilot test study instruments, train data collection teams, employ specific sampling strategies (e.g., recruiting patients diagnosed within the last six months), and utilize data triangulation, including mixed methods and comparison to earlier studies where possible. Additionally, data collectors need to be creative in helping patients recall key dates, e.g. with use of local or national events, and/or use clinical records to verify patient-reported dates, where feasible.

Conceptually, incorporating patient cost surveys into journey analyses is another opportunity for a comprehensive understanding of patient care barriers, particularly where these include incurred costs for all healthcare provider visits. Cost surveys are often conducted by economists with a primary focus on cost analysis, and rarely provide detailed insights into the nuances of patient journeys. Conversely, patient journeys analyses rarely include cost data.

This disconnect between cost analysis and journey mapping presents a significant gap in the literature and integrating both aspects could provide a more holistic view of patient experiences and economic impacts. To address this gap, we recommend that future patient cost surveys be designed to enable rigorous analysis of patient journeys, and vice versa. Such an approach would facilitate a better understanding of both the financial and experiential data points of patient care-seeking behaviours. Ultimately, this dual-focus approach could significantly enhance the quality of insights derived from patient surveys, leading to improved patient-centric service delivery and better-informed healthcare policy decisions.

## Conclusions

Our findings underscore the critical need for better approaches to compare patient journeys and develop interventions, emphasizing the importance of clear guidelines and accessible tools to facilitate this analysis. This work lays the groundwork for future efforts to enhance access, quality, and equity in healthcare service provision.

## Supplementary materials

### Supplement 1 - Methodological Review Search Strategy

CINAHL

No field selected; Suggest subject terms checked

Total: 519 [6 Apr 29], update TB: 19, May 05

| Data points | Key words |
| --- | --- |
| Patient journey | (TI ( "Patient pathway analys*" OR PPA OR "treatment delay" OR "health-seeking" OR "healthcare-seeking" "health-seeking delay" OR "care seeking" OR "cascade of care" OR "access to care" OR "healthcare delivery" OR "diagnostic delay" OR "care seeking delay" OR "barriers and facilitators to care" OR "missed opportunit*" OR "care pathway*" ) OR AB ( "Patient pathway analys*" OR PPA OR "treatment delay" OR "health-seeking" OR "health-seeking delay" OR "care seeking" OR "cascade of care" OR "access to care" OR "healthcare delivery" OR "diagnostic delay" OR "care seeking delay" OR "barriers and facilitators to care" OR "missed opportunit*" OR "care pathway*" ) OR (MH "Diagnosis, Delayed") ) |
| Tuberculosis OR other pulmonary diseases OR selected infectious diseases | ((MH "Tuberculosis, Pulmonary") OR TI (tb OR tuberculosis) OR AB(tb OR tuberculosis) OR (MH "Mycobacterium Tuberculosis")) OR (MH "respiratory tract diseases") OR (MH "respiratory disease") OR (MH "respiratory infection") OR (MH "malaria") OR TI (malaria OR HIV OR covid) OR AB (malaria OR HIV OR covid) OR (MH "human immunodeficiency virus") OR (MH "sars-cov-2") OR (MH "covid-19")) |

PubMed

Total: 1631 [1625 apr 29], update TB: 100 May 05

Health seeking pathways (May 8):

| Data points | Key words |
| --- | --- |
| Patient journey analysis | "Patient pathway analys*" [tw] OR PPA [tw] OR "treatment delay" [tw] OR "healthcare-seeking" [tw] OR "health-seeking delay" [tw] OR "care seeking" [tw] OR "cascade of care" [tw] OR "access to care" [tw] OR "healthcare delivery" [tw] OR "diagnostic delay" [tw] OR "pathway analysis" [tw] OR "care seeking delay" [tw] OR "barriers and facilitators to care" [tw] OR "missed opportunit*" [tw] OR "care pathway*" [tw] OR "delayed diagnosis" [MeSH] OR "critical pathways" [MeSH] OR "patient acceptance of health care"[MeSH] |
| Tuberculosis | Tuberculosis, Pulmonary [Mesh] OR tb[tw] OR tuberculosis[tw] OR Mycobacterium tuberculosis[Mesh] |
| Other pulmonary | "respiratory tract diseases"[MeSH Terms] OR "respiration |
| diseases and relevant infectious disease | disorders"[MeSH Terms] OR respiratory disease[Text Word] OR "respiratory infection" OR "malaria"[MeSH Terms] OR malaria [tw] OR "hiv"[MeSH Terms] OR HIV[tw] OR "human immunodeficiency virus" [tw] "sars-cov-2"[MeSH Terms] OR "covid-19"[MeSH Terms] OR covid [tw] |

Web of Science

TS search; no filter

Total: 1336 (3518 apr 29), update TB: 58, May 05

| Data points | Key words |
| --- | --- |
| Patient journey analysis | TS=( "Patient pathway analys*" OR PPA OR "treatment delay" OR "healthcare-seeking" OR "health-seeking delay" OR "care seeking" OR "cascade of care" OR "access to care" OR "healthcare delivery" OR "diagnostic delay" OR "care seeking delay" OR "barriers and facilitators to care" OR "missed opportunit*" OR "care pathway*" ) |
| Tuberculosis | TS=(tb OR tuberculosis) |
| Other pulmonary diseases and relevant infectious disease | TS=("respiratory tract diseases" OR "respiration disorders" OR "respiratory disease" OR "respiratory infection" OR "malaria" OR HIV OR "human immunodeficiency virus" OR "sars-cov-2" OR "covid-19" OR covid) |

Global Health via Ovid

Total: 932 (2798 apr 29), update TB: 100 May 05

No filter

| Data points | Key words |
| --- | --- |
| Patient journey analysis | ( "Patient pathway analys*" OR PPA OR "treatment delay" OR "healthcare-seeking" OR "health-seeking delay" OR "care seeking" OR "cascade of care" OR "access to care" OR "healthcare delivery" OR "diagnostic delay" OR "care seeking delay" OR "barriers and facilitators to care" OR "missed opportunit*" OR "care pathway*" ).mp. |
| Tuberculosis | (tb OR tuberculosis).mp. |
|  | ("respiratory tract diseases" or "respiration disorders" or "respiratory disease" or "respiratory infection" or malaria or HIV or "human immunodeficiency virus" or "sars-cov-2" or covid-19 or covid).mp. |

### Supplement 2 - Data extraction form

#### General information

|  |  |
| --- | --- |
| First author last name | [Open text field] |
| Year of publication | [Open text field] |
| Country/setting<br>as specific as possible | [Open text field] |
| Disease type<br>TB, HIV, malaria, etc. | [Open text field] |
| Study design/Data sources<br>e.g. population-based surveys, patient<br>interviews | [Open text field] |
| Quantitative/Qualitative/Mixed | [Open text field] |
| Total sample size<br>Indicate sex of participants if available | [Open text field] |

#### Patient healthcare-seeking journey data points included

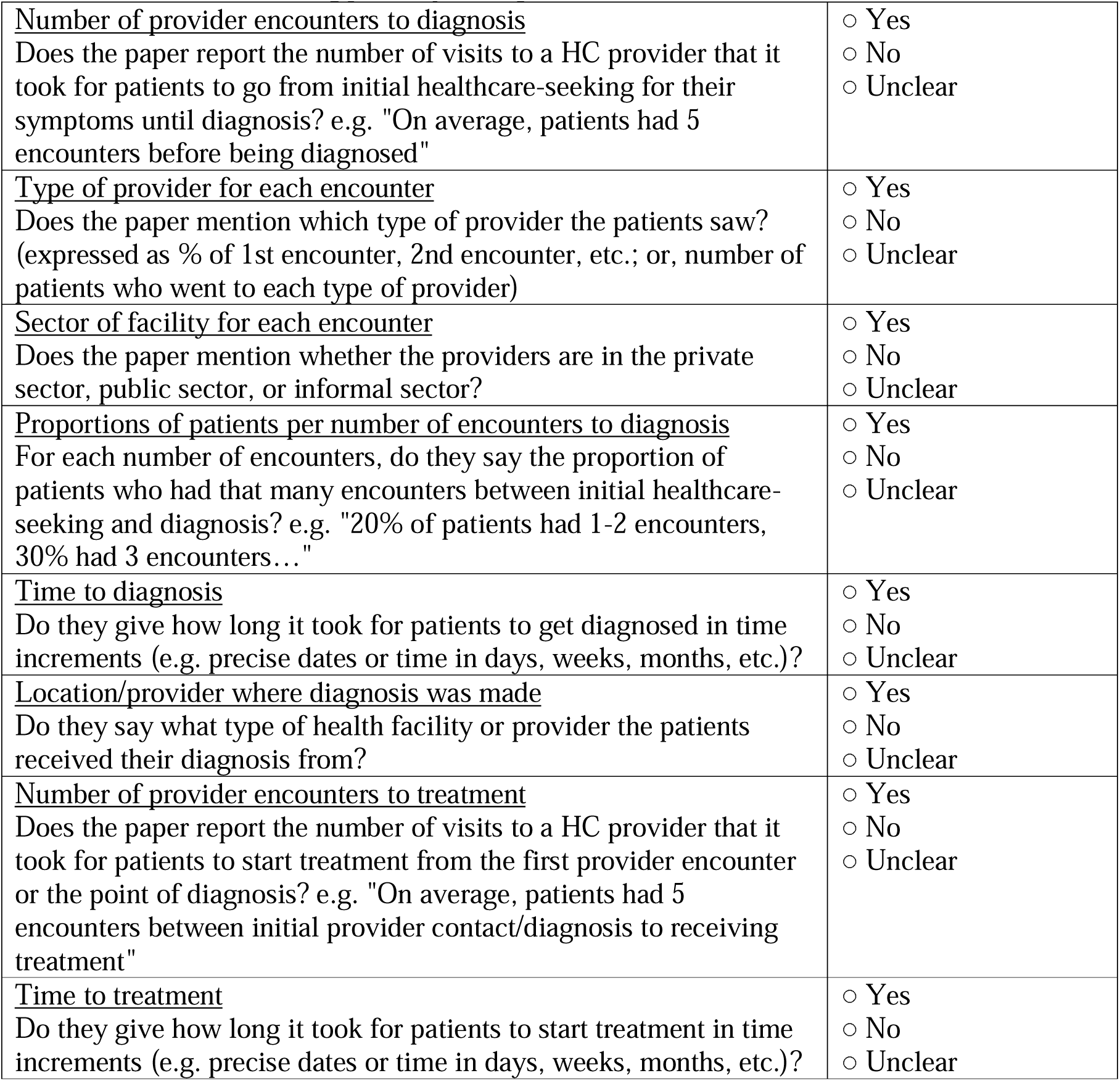
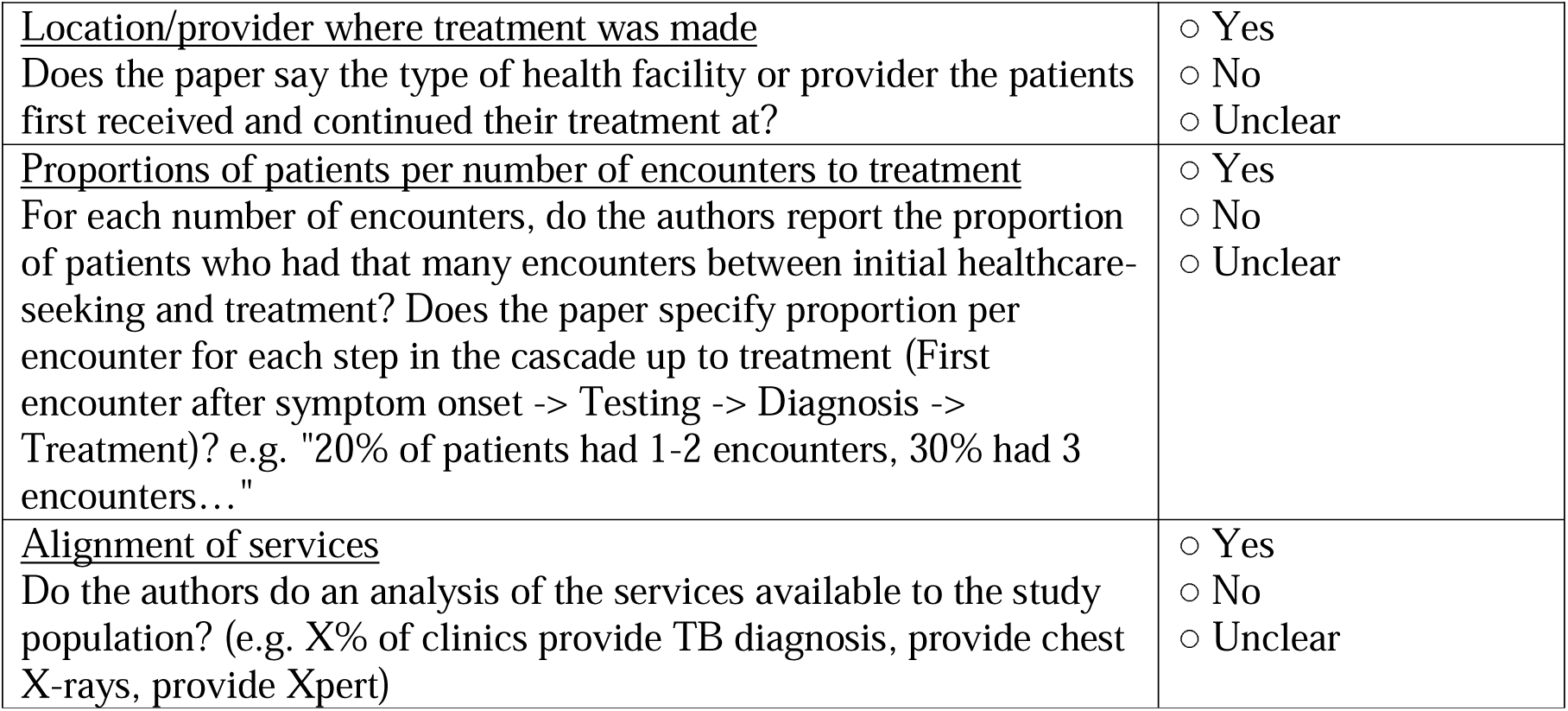

|  |  |
| --- | --- |
| <u>Number of provider encounters to diagnosis</u><br>Does the paper report the number of visits to a HC provider that it took for patients to go from initial healthcare-seeking for their symptoms until diagnosis? e.g. "On average, patients had 5 encounters before being diagnosed" | ☐ Yes<br>☐ No<br>☐ Unclear |
| <u>Type of provider for each encounter</u><br>Does the paper mention which type of provider the patients saw? (expressed as % of 1st encounter, 2nd encounter, etc.; or, number of patients who went to each type of provider) | ☐ Yes<br>☐ No<br>☐ Unclear |
| <u>Sector of facility for each encounter</u><br>Does the paper mention whether the providers are in the private sector, public sector, or informal sector? | ☐ Yes<br>☐ No<br>☐ Unclear |
| <u>Proportions of patients per number of encounters to diagnosis</u><br>For each number of encounters, do they say the proportion of patients who had that many encounters between initial healthcare-seeking and diagnosis? e.g. "20% of patients had 1-2 encounters, 30% had 3 encounters..." | ☐ Yes<br>☐ No<br>☐ Unclear |
| <u>Time to diagnosis</u><br>Do they give how long it took for patients to get diagnosed in time increments (e.g. precise dates or time in days, weeks, months, etc.)? | ☐ Yes<br>☐ No<br>☐ Unclear |
| <u>Location/provider where diagnosis was made</u><br>Do they say what type of health facility or provider the patients received their diagnosis from? | ☐ Yes<br>☐ No<br>☐ Unclear |
| <u>Number of provider encounters to treatment</u><br>Does the paper report the number of visits to a HC provider that it took for patients to start treatment from the first provider encounter or the point of diagnosis? e.g. "On average, patients had 5 encounters between initial provider contact/diagnosis to receiving treatment" | ☐ Yes<br>☐ No<br>☐ Unclear |
| <u>Time to treatment</u><br>Do they give how long it took for patients to start treatment in time increments (e.g. precise dates or time in days, weeks, months, etc.)? | ☐ Yes<br>☐ No<br>☐ Unclear |
| <u>Location/provider where treatment was made</u><br>Does the paper say the type of health facility or provider the patients first received and continued their treatment at? | ☐ Yes<br>☐ No<br>☐ Unclear |
| <u>Proportions of patients per number of encounters to treatment</u><br>For each number of encounters, do the authors report the proportion of patients who had that many encounters between initial healthcare-seeking and treatment? Does the paper specify proportion per encounter for each step in the cascade up to treatment (First encounter after symptom onset -> Testing -> Diagnosis -> Treatment)? e.g. "20% of patients had 1-2 encounters, 30% had 3 encounters..." | ☐ Yes<br>☐ No<br>☐ Unclear |
| <u>Alignment of services</u><br>Do the authors do an analysis of the services available to the study population? (e.g. X% of clinics provide TB diagnosis, provide chest X-rays, provide Xpert) | ☐ Yes<br>☐ No<br>☐ Unclear |

#### Journey methodology & analysis

|  |  |
| --- | --- |
| <u>Name or description of journey methodology (type of chart)</u><br>e.g. Hansen-Chin, flowchart, Sankey chart, etc. Indicate if author's own methodology or replication of previous one. If author's own, indicate main components. | [Open text field] |
| <u>Summary of analysis</u><br>Description of statistical analysis | [Open text field] |
| <u>Outcome variables or major themes</u> | [Open text field] |
| <u>Strengths</u><br>If authors tried to limit recall bias, indicate how. | [Open text field] |
| <u>Limitations</u> | [Open text field] |
| <u>Variables of interest or minor themes</u><br>Indicate the specific outcome that the variable is in reference to (cost, delay, access, etc.) | [Open text field] |

Outcome measures

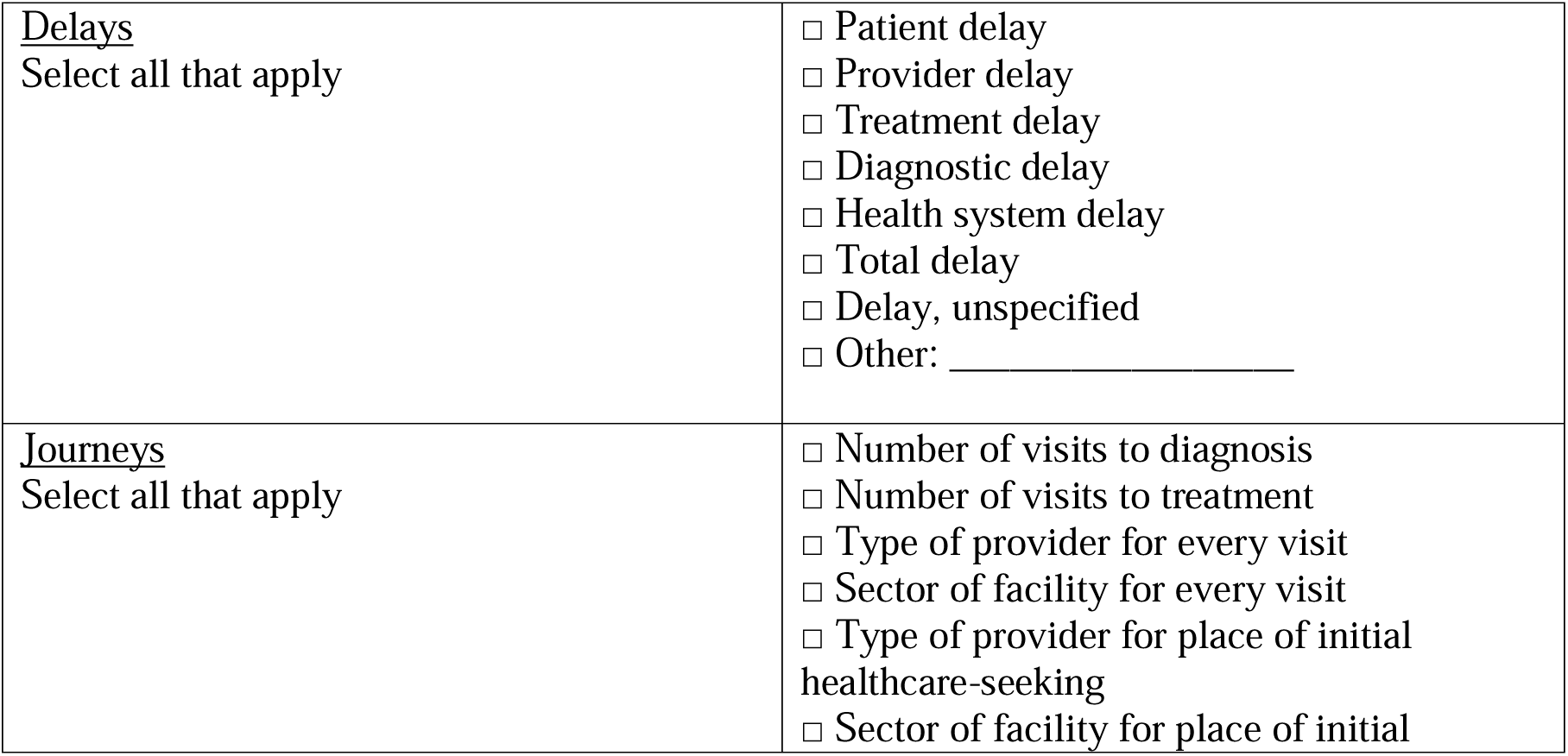
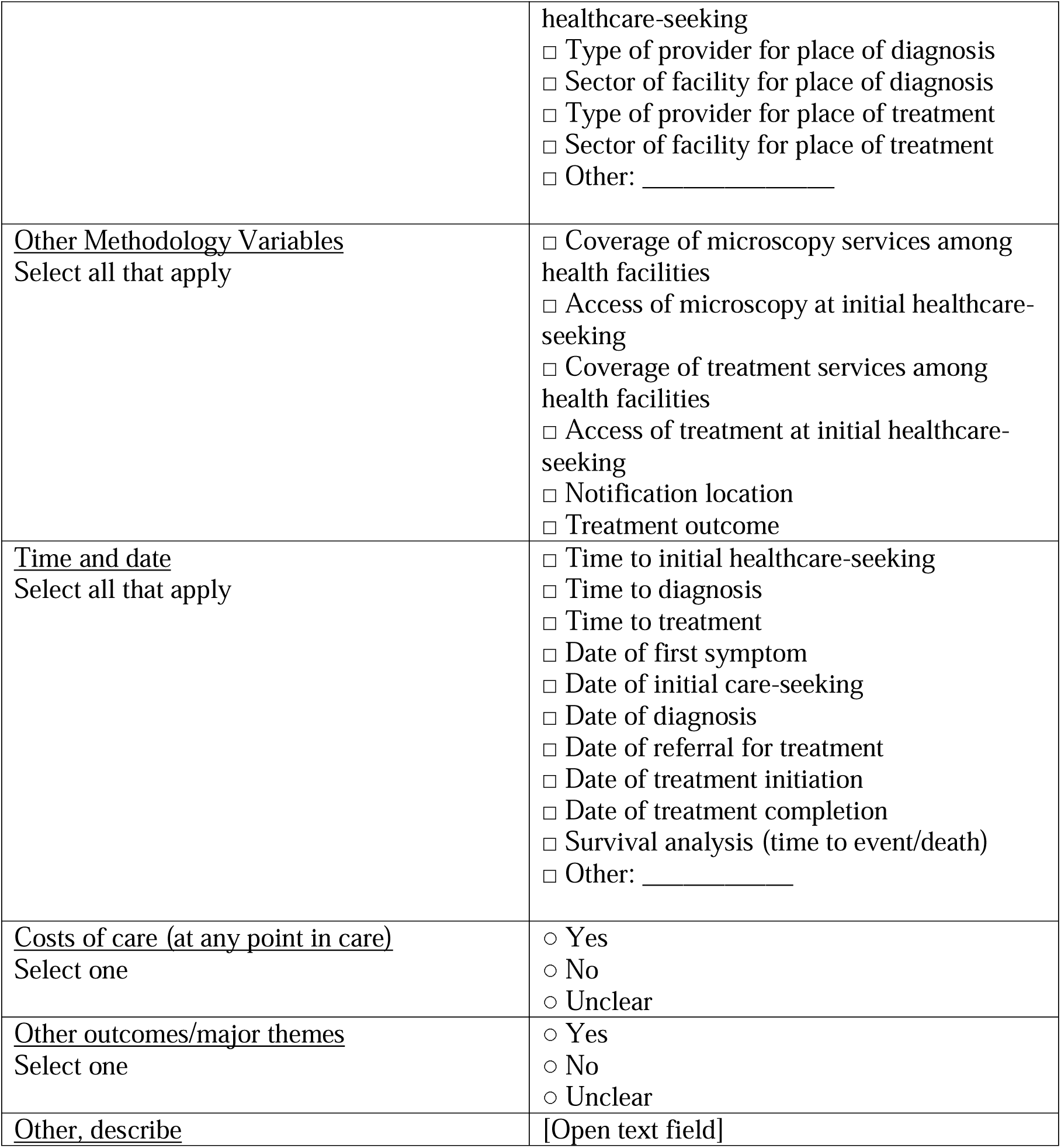

Other variables

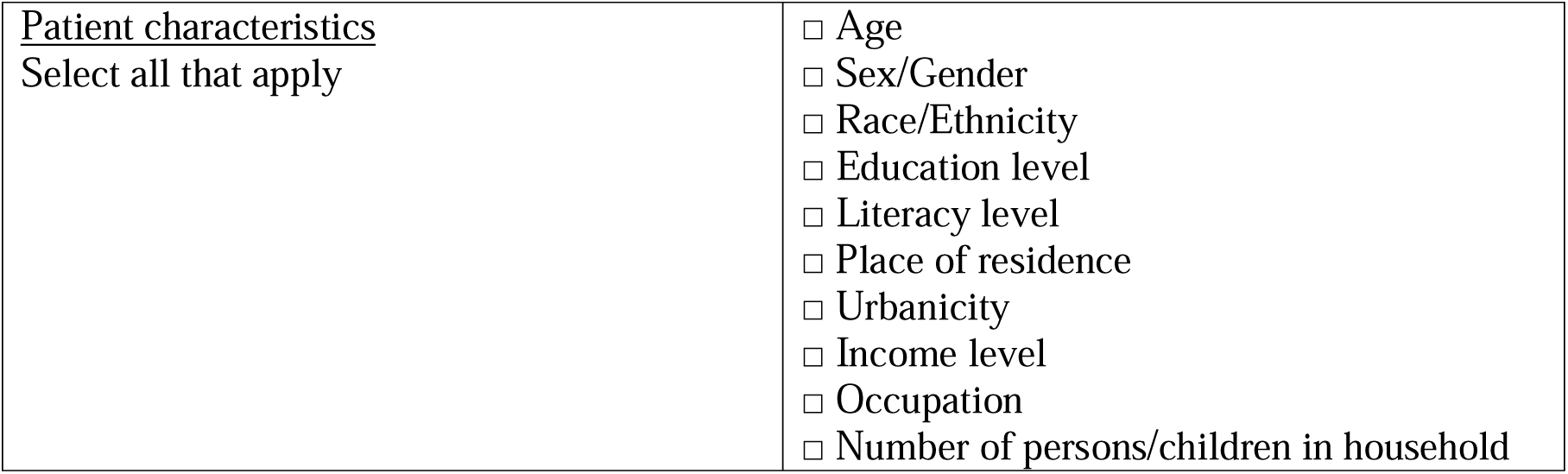
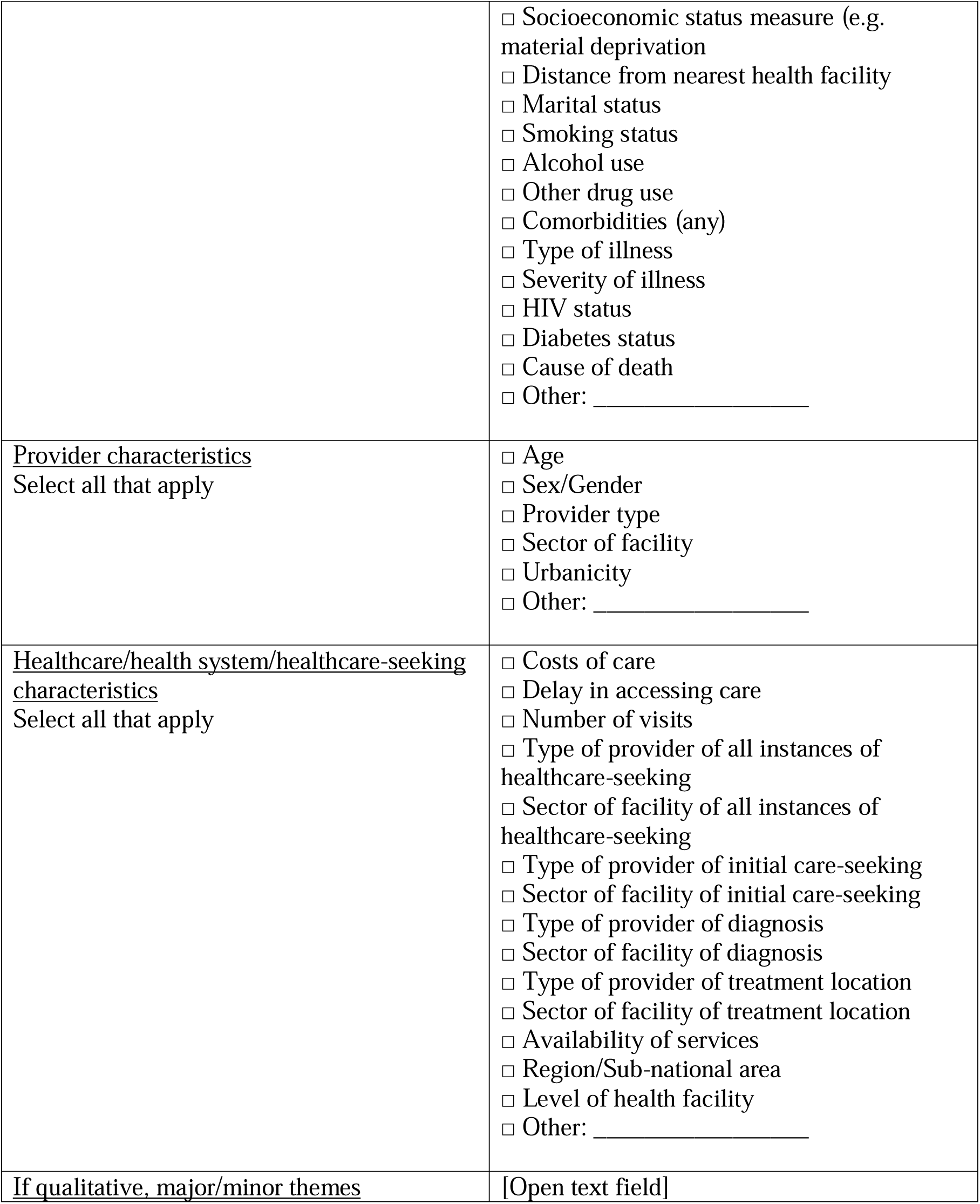

### Supplement 3 – Quality Assessment Electronic Forms

#### General information

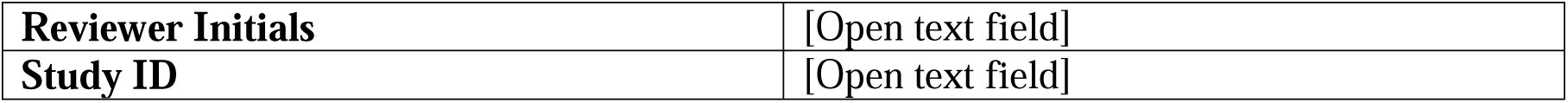
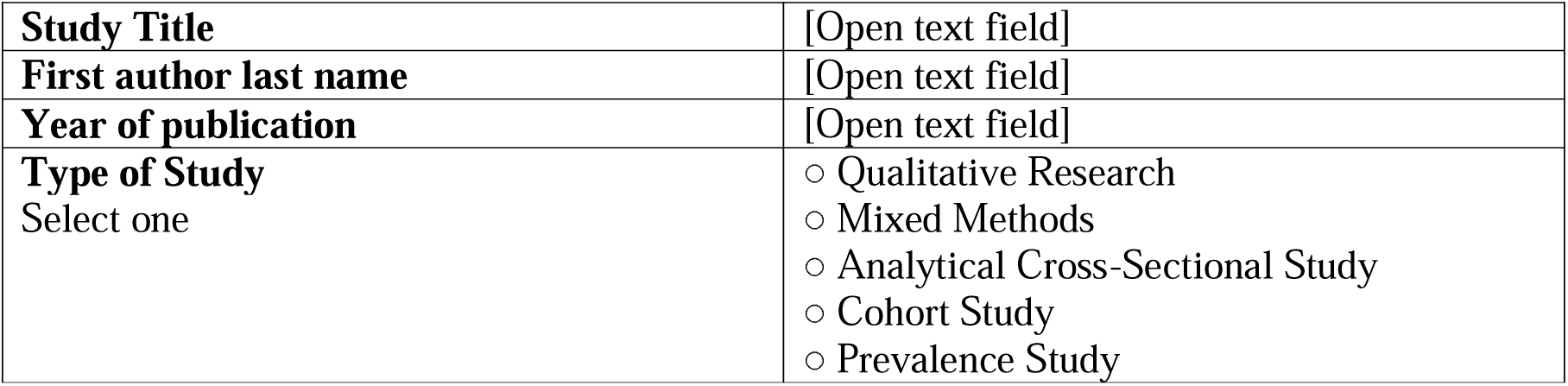

#### Qualitative Research Quality Assessment

Source: https://jbi.global/sites/default/files/2021-10/Checklist_for_Qualitative_Research.docx

Discussion of critical appraisal criteria are added before each question for context.

|  |  |
| --- | --- |
| **1. Is there congruity between the stated philosophical perspective and the research methodology**<br>Does the report clearly state the philosophical or theoretical premises on which the study is based? Does the report clearly state the methodological approach adopted on which the study is based? Is there congruence between the two? For example:<br>A report may state that the study adopted a critical perspective and participatory action research methodology was followed. Here there is congruence between a critical view (focusing on knowledge arising out of critique, action and reflection) and action research (an approach that focuses on firstly working with groups to reflect on issues or practices, then considering how they could be different; then acting to create a change; and finally identifying new knowledge arising out of the action taken). However, a report may state that the study adopted an interpretive perspective and used survey methodology. Here there is incongruence between an interpretive view (focusing on knowledge arising out of studying what phenomena mean to individuals or groups) and surveys (an approach that focuses on asking standard questions to a defined study population); a report may state that the study was qualitative or used qualitative methodology (such statements do not demonstrate rigour in design) or make no statement on philosophical orientation or methodology. | ☐ Yes<br>☐ No<br>☐ Unclear<br>☐ Not applicable |
| **2. Is there congruity between the research methodology and the research question or objectives?**<br>Is the study methodology appropriate for addressing the research question? For example:<br>A report may state that the research question was to seek understandings of the meaning of pain in a group of people with | ☐ Yes<br>☐ No<br>☐ Unclear<br>☐ Not applicable |
| rheumatoid arthritis and that a phenomenological approach was taken. Here, there is congruity between this question and the methodology. A report may state that the research question was to establish the effects of counselling on the severity of pain experience and that an ethnographic approach was pursued. A question that tries to establish cause-and effect cannot be addressed by using an ethnographic approach (as ethnography sets out to develop understandings of cultural practices) and thus, this would be incongruent. |  |
| **3. Is there congruity between the research methodology and the methods used to collect data**<br>Are the data collection methods appropriate to the methodology?<br>For example:<br>A report may state that the study pursued a phenomenological approach and data was collected through phenomenological interviews. There is congruence between the methodology and data collection; a report may state that the study pursued a phenomenological approach and data was collected through a postal questionnaire. There is incongruence between the methodology and data collection here as phenomenology seeks to elicit rich descriptions of the experience of a phenomena that cannot be achieved through seeking written responses to standardized questions. | ☐ Yes<br>☐ No<br>☐ Unclear<br>☐ Not applicable |
| **4. Is there congruity between the research methodology and the representation and analysis of data?**<br>Are the data analyzed and represented in ways that are congruent with the stated methodological position? For example:<br>A report may state that the study pursued a phenomenological approach to explore people's experience of grief by asking participants to describe their experiences of grief. If the text generated from asking these questions is searched to establish the meaning of grief to participants, and the meanings of all participants are included in the report findings, then this represents congruity; the same report may, however, focus only on those meanings that were common to all participants and discard single reported meanings. This would not be appropriate in phenomenological work. | ☐ Yes<br>☐ No<br>☐ Unclear<br>☐ Not applicable |
| **5. Is there congruence between the research methodology and the interpretation of results?**<br>Are the results interpreted in ways that are appropriate to the methodology? For example:<br>A report may state that the study pursued a phenomenological approach to explore people's experience of facial disfigurement and | ☐ Yes<br>☐ No<br>☐ Unclear<br>☐ Not applicable |
| the results are used to inform practitioners about accommodating individual differences in care. There is congruence between the methodology and this approach to interpretation; a report may state that the study pursued a phenomenological approach to explore people's experience of facial disfigurement and the results are used to generate practice checklists for assessment. There is incongruence between the methodology and this approach to interpretation as phenomenology seeks to understand the meaning of a phenomenon for the study participants and cannot be interpreted to suggest that this can be generalized to total populations to a degree where standardized assessments will have relevance across a population. |  |
| **6. Is there a statement locating the researcher culturally or theoretically?**<br>Are the beliefs and values, and their potential influence on the study declared? For example:<br>The researcher plays a substantial role in the qualitative research process and it is important, in appraising evidence that is generated in this way, to know the researcher's cultural and theoretical orientation. A high quality report will include a statement that clarifies this. | ☐ Yes<br>☐ No<br>☐ Unclear<br>☐ Not applicable |
| **7. Is the influence of the researcher on the research, and vice-versa, addressed?**<br>Is the potential for the researcher to influence the study and for the potential of the research process itself to influence the researcher and her/his interpretations acknowledged and addressed? For example:<br>Is the relationship between the researcher and the study participants addressed? Does the researcher critically examine her/his own role and potential influence during data collection? Is it reported how the researcher responded to events that arose during the study? | ☐ Yes<br>☐ No<br>☐ Unclear<br>☐ Not applicable |
| **8. Are participants, and their voices, adequately represented?**<br>Generally, reports should provide illustrations from the data to show the basis of their conclusions and to ensure that participants are represented in the report. | ☐ Yes<br>☐ No<br>☐ Unclear<br>☐ Not applicable |
| **9. Is the research ethical according to current criteria or, for recent studies, and is there evidence of ethical approval by an appropriate body?**<br>A statement on the ethical approval process followed should be in the report. | ☐ Yes<br>☐ No<br>☐ Unclear<br>☐ Not applicable |
| **10. Do the conclusions drawn in the research report flow from the analysis, or interpretation, of the data?**<br>This criterion concerns the relationship between the findings reported and the views or words of study participants. In appraising | ☐ Yes<br>☐ No<br>☐ Unclear<br>☐ Not applicable |
| a paper, appraisers seek to satisfy themselves that the conclusions drawn by the research are based on the data collected; data being the text generated through observation, interviews or other processes. |  |
| **Overall appraisal** | ☐ Low quality<br>☐ Medium quality<br>☐ High quality<br>☐ Seek further info |
| **Comments** | [Open text field] |

#### Mixed Methods Quality Assessment

Adapted from the Mixed Methods Appraisal Tool. Citation: Hong QN, Pluye P, Fàbregues S, Bartlett G, Boardman F, Cargo M, Dagenais P, Gagnon M-P, Griffiths F, Nicolau B, O’Cathain A, Rousseau M-C, Vedel I. Mixed Methods Appraisal Tool (MMAT), version 2018. Registration of Copyright (#1148552), Canadian Intellectual Property Office, Industry Canada.

|  |  |
| --- | --- |
| 1. Are there clear research questions? | ☐ Yes<br>☐ No<br>☐ Unclear<br>☐ Not applicable |
| 2. Do the collected data allow the research questions to be addressed? | ☐ Yes<br>☐ No<br>☐ Unclear<br>☐ Not applicable |
| **Part 1: Assessment of the Qualitative Aspects of the Study** |  |
| Is the qualitative approach appropriate to answer the research question? | ☐ Yes<br>☐ No<br>☐ Unclear<br>☐ Not applicable |
| Are the qualitative data collection methods adequate to address the research question? | ☐ Yes<br>☐ No<br>☐ Unclear<br>☐ Not applicable |
| Are the findings adequately derived from the data? | ☐ Yes<br>☐ No<br>☐ Unclear<br>☐ Not applicable |
| Is the interpretation of results sufficiently substantiated by data? | ☐ Yes<br>☐ No<br>☐ Unclear<br>☐ Not applicable |
| Is there coherence between qualitative data sources, collection, analysis, and interpretation? | ☐ Yes<br>☐ No<br>☐ Unclear<br>☐ Not applicable |
| **Part 2: Assessment of the Quantitative Aspects of the Study** |  |
| Is the sampling strategy relevant to address the research question? | ☐ Yes<br>☐ No<br>☐ Unclear |
|  | ○ Not applicable |
| Is the sample representative of the target population? | ○ Yes<br>○ No<br>○ Unclear<br>○ Not applicable |
| Are the measurements appropriate? | ○ Yes<br>○ No<br>○ Unclear<br>○ Not applicable |
| Is the risk of nonresponse bias low? | ○ Yes<br>○ No<br>○ Unclear<br>○ Not applicable |
| **Is the statistical analysis appropriate to answer the research question?** | ○ Yes<br>○ No<br>○ Unclear<br>○ Not applicable |
| **Part 3: Assessment of the Mixed Methods Aspects of the Study** |  |
| Is there an adequate rationale for using a mixed methods design to address the research question? | ○ Yes<br>○ No<br>○ Unclear<br>○ Not applicable |
| Are the different components of the study effectively integrated to answer the research question? | ○ Yes<br>○ No<br>○ Unclear<br>○ Not applicable |
| Are the outputs of the integration of qualitative and quantitative components adequately interpreted? | ○ Yes<br>○ No<br>○ Unclear<br>○ Not applicable |
| Are divergences and inconsistencies between quantitative and qualitative results adequately addressed? | ○ Yes<br>○ No<br>○ Unclear<br>○ Not applicable |
| Do the different components of the study adhere to the quality criteria of each tradition of the methods involved? | ○ Yes<br>○ No<br>○ Unclear<br>○ Not applicable |
| Overall appraisal | ○ Low quality<br>○ Medium quality<br>○ High quality<br>○ Seek further info |
| Comments | [Open text field] |

#### Analytical Cross-Sectional Quality Assessment

Source: https://jbi.global/sites/default/files/2021-10/Checklist_for_Analytical_Cross_Sectional_Studies.docx

Discussion of critical appraisal criteria are added before each question for context.

|  |  |
| --- | --- |
| **1. Were the criteria for inclusion in the sample clearly defined?**<br>The authors should provide clear inclusion and exclusion criteria that they developed prior to recruitment of the study participants. The inclusion/exclusion criteria should be specified (e.g., risk, stage of disease progression) with sufficient detail and all the necessary information critical to the study. | ☐ Yes<br>☐ No<br>☐ Unclear<br>☐ Not applicable |
| **2. Were the study subjects and the setting described in detail?**<br>The study sample should be described in sufficient detail so that other researchers can determine if it is comparable to the population of interest to them. The authors should provide a clear description of the population from which the study participants were selected or recruited, including demographics, location, and time period. | ☐ Yes<br>☐ No<br>☐ Unclear<br>☐ Not applicable |
| **3. Was the exposure measured in a valid and reliable way?**<br>The study should clearly describe the method of measurement of exposure. Assessing validity requires that a 'gold standard' is available to which the measure can be compared. The validity of exposure measurement usually relates to whether a current measure is appropriate or whether a measure of past exposure is needed.<br>Reliability refers to the processes included in an epidemiological study to check repeatability of measurements of the exposures. These usually include intra-observer reliability and inter-observer reliability. | ☐ Yes<br>☐ No<br>☐ Unclear<br>☐ Not applicable |
| **4. Were objective, standard criteria used for measurement of the condition?**<br>It is useful to determine if patients were included in the study based on either a specified diagnosis or definition. This is more likely to decrease the risk of bias. Characteristics are another useful approach to matching groups, and studies that did not use specified diagnostic methods or definitions should provide evidence on matching by key characteristics. | ☐ Yes<br>☐ No<br>☐ Unclear<br>☐ Not applicable |
| **5. Were confounding factors identified?**<br>Confounding has occurred where the estimated intervention exposure effect is biased by the presence of some difference between the comparison groups (apart from the exposure investigated/of interest). Typical confounders include baseline characteristics, prognostic factors, or concomitant exposures (e.g. smoking). A confounder is a difference between the comparison groups and it influences the direction of the study results. A high quality study at the level of cohort design will identify the potential confounders and measure them (where possible). This is difficult for studies where behavioural, attitudinal or lifestyle factors may impact on the results. | ☐ Yes<br>☐ No<br>☐ Unclear<br>☐ Not applicable |
| **6. Were strategies to deal with confounding factors stated?**<br>Strategies to deal with effects of confounding factors may be dealt within the study design or in data analysis. By matching or stratifying sampling of participants, effects of confounding factors can be adjusted for. When dealing with adjustment in data analysis, | ☐ Yes<br>☐ No<br>☐ Unclear<br>☐ Not applicable |
| assess the statistics used in the study. Most will be some form of multivariate regression analysis to account for the confounding factors measured. |  |
| **7. Were the outcomes measured in a valid and reliable way?**<br>Read the methods section of the paper. If for e.g. lung cancer is assessed based on existing definitions or diagnostic criteria, then the answer to this question is likely to be yes. If lung cancer is assessed using observer reported, or self-reported scales, the risk of over- or under-reporting is increased, and objectivity is compromised. Importantly, determine if the measurement tools used were validated instruments as this has a significant impact on outcome assessment validity.<br>Having established the objectivity of the outcome measurement (e.g. lung cancer) instrument, it's important to establish how the measurement was conducted. Were those involved in collecting data trained or educated in the use of the instrument/s? (e.g. radiographers). If there was more than one data collector, were they similar in terms of level of education, clinical or research experience, or level of responsibility in the piece of research being appraised? | ☐ Yes<br>☐ No<br>☐ Unclear<br>☐ Not applicable |
| **8. Was appropriate statistical analysis used?**<br>As with any consideration of statistical analysis, consideration should be given to whether there was a more appropriate alternate statistical method that could have been used. The methods section should be detailed enough for reviewers to identify which analytical techniques were used (in particular, regression or stratification) and how specific confounders were measured.<br>For studies utilizing regression analysis, it is useful to identify if the study identified which variables were included and how they related to the outcome. If stratification was the analytical approach used, were the strata of analysis defined by the specified variables? Additionally, it is also important to assess the appropriateness of the analytical strategy in terms of the assumptions associated with the approach as differing methods of analysis are based on differing assumptions about the data and how it will respond. | ☐ Yes<br>☐ No<br>☐ Unclear<br>☐ Not applicable |
| **Overall appraisal** | ☐ Low quality<br>☐ Medium quality<br>☐ High quality<br>☐ Seek further info |
| **Comments** | [Open text field] |

#### Cohort Study Quality Assessment

Source: https://jbi.global/sites/default/files/2021-10/Checklist_for_Cohort_Studies.docx

Discussion of critical appraisal criteria are added before each question for context.

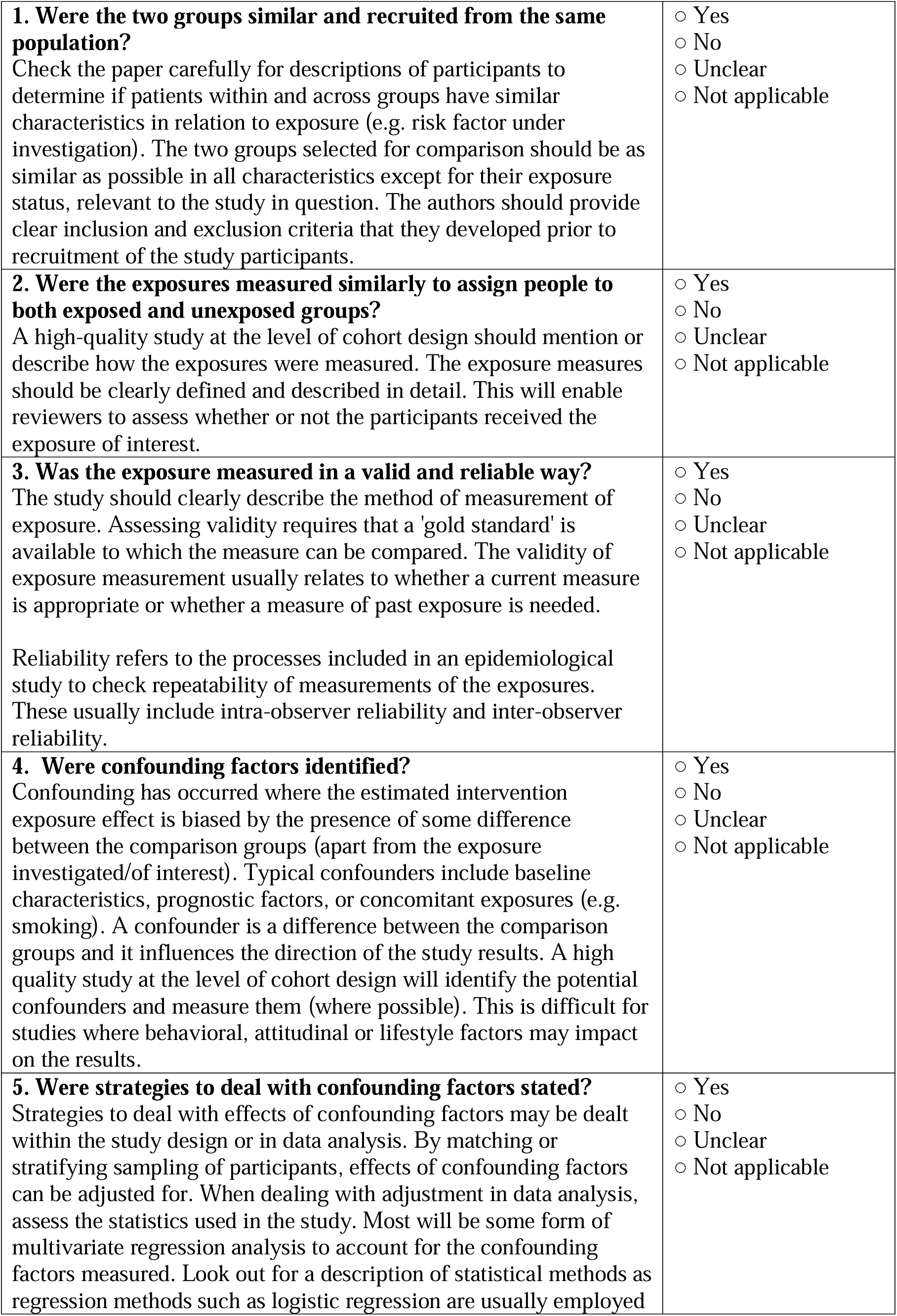
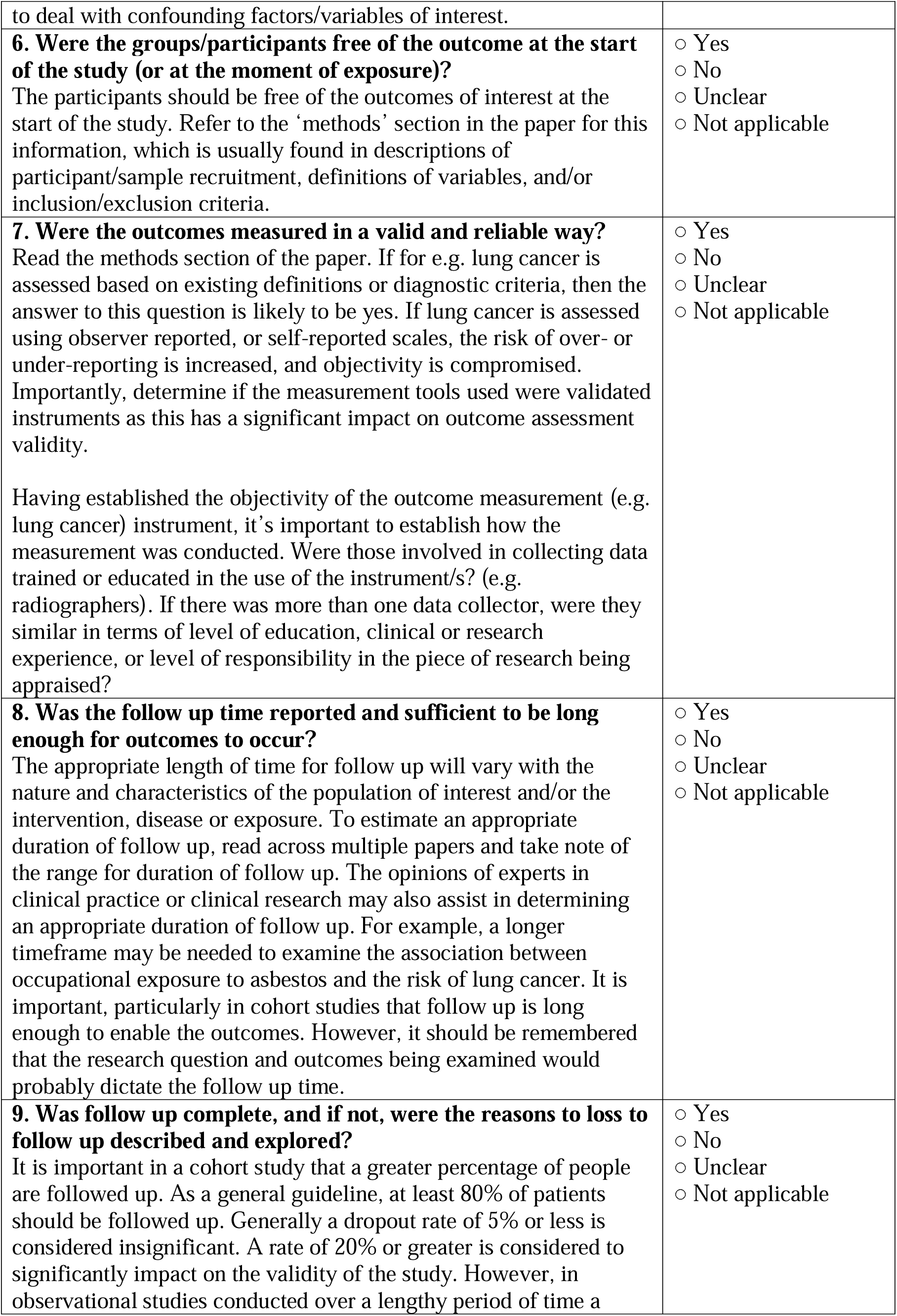
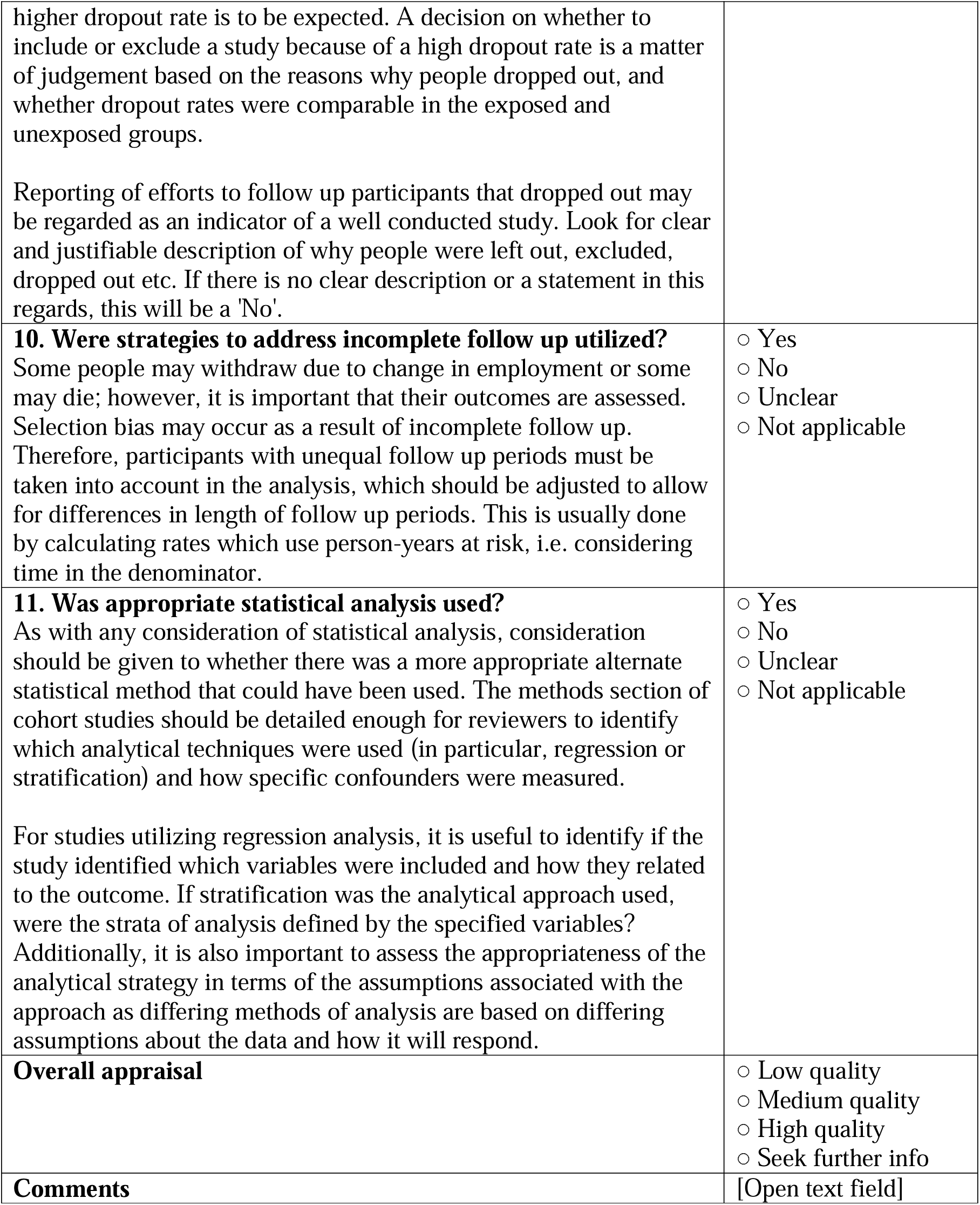

#### Prevalence Study Quality Assessment

Source: https://jbi.global/sites/default/files/2021-10/Checklist_for_Prevalence_Studies.docx

Discussion of critical appraisal criteria are added before each question for context.

|  |  |
| --- | --- |
| **1. Was the sample frame appropriate to address the target population?** | ☐ Yes<br>☐ No |
| **This question relies upon knowledge of the broader characteristics of the population of interest and the geographical area.** If the study is of women with breast cancer, knowledge of at least the characteristics, demographics and medical history is needed. The term “target population” should not be taken to infer every individual from everywhere or with similar disease or exposure characteristics. Instead, give consideration to specific population characteristics in the study, including age range, gender, morbidities, medications, and other potentially influential factors. For example, a sample frame may not be appropriate to address the target population if a certain group has been used (such as those working for one organisation, or one profession) and the results then inferred to the target population (i.e. working adults). A sample frame may be appropriate when it includes almost all the members of the target population (i.e. a census, or a complete list of participants or complete registry data). | ○ Unclear<br>○ Not applicable |
| **2. Were study participants recruited in an appropriate way?** Studies may report random sampling from a population, and the methods section should report how sampling was performed. Random probabilistic sampling from a defined subset of the population (sample frame) should be employed in most cases, however, random probabilistic sampling is not needed when everyone in the sampling frame will be included/analysed. For example, reporting on all the data from a good census is appropriate as a good census will identify everybody. When using cluster sampling, such as a random sample of villages within a region, the methods need to be clearly stated as the precision of the final prevalence estimate incorporates the clustering effect. Convenience samples, such as a street survey or interviewing lots of people at a public gatherings are not considered to provide a representative sample of the base population. | ○ Yes<br>○ No<br>○ Unclear<br>○ Not applicable |
| **3. Was the sample size adequate?** The larger the sample, the narrower will be the confidence interval around the prevalence estimate, making the results more precise. An adequate sample size is important to ensure good precision of the final estimate. Ideally we are looking for evidence that the authors conducted a sample size calculation to determine an adequate sample size. This will estimate how many subjects are needed to produce a reliable estimate of the measure(s) of interest. For conditions with a low prevalence, a larger sample size is needed. Also consider sample sizes for subgroup (or characteristics) analyses, and whether these are appropriate. Sometimes, the study will be large enough (as in large national surveys) whereby a sample size calculation is not required. In these cases, sample size can be considered adequate. When there is no sample size calculation and it is not a large national survey, the reviewers may consider conducting their own sample size analysis. | ○ Yes<br>○ No<br>○ Unclear<br>○ Not applicable |
| **4. Were the study subjects and setting described in detail?**<br>Certain diseases or conditions vary in prevalence across different geographic regions and populations (e.g. Women vs. Men, sociodemographic variables between countries). The study sample should be described in sufficient detail so that other researchers can determine if it is comparable to the population of interest to them. | ☐ Yes<br>☐ No<br>☐ Unclear<br>☐ Not applicable |
| **5. Was data analysis conducted with sufficient coverage of the identified sample?**<br>Coverage bias can occur when not all subgroups of the identified sample respond at the same rate. For instance, you may have a very high response rate overall for your study, but the response rate for a certain subgroup (i.e. older adults) may be quite low. | ☐ Yes<br>☐ No<br>☐ Unclear<br>☐ Not applicable |
| **6. Were valid methods used for the identification of the condition?**<br>Here we are looking for measurement or classification bias. Many health problems are not easily diagnosed or defined and some measures may not be capable of including or excluding appropriate levels or stages of the health problem. If the outcomes were assessed based on existing definitions or diagnostic criteria, then the answer to this question is likely to be yes. If the outcomes were assessed using observer reported, or self-reported scales, the risk of over- or under-reporting is increased, and objectivity is compromised. Importantly, determine if the measurement tools used were validated instruments as this has a significant impact on outcome assessment validity. | ☐ Yes<br>☐ No<br>☐ Unclear<br>☐ Not applicable |
| **7. Was the condition measured in a standard, reliable way for all participants?**<br>Considerable judgment is required to determine the presence of some health outcomes. Having established the validity of the outcome measurement instrument (see item 6 of this scale), it is important to establish how the measurement was conducted. Were those involved in collecting data trained or educated in the use of the instrument/s? If there was more than one data collector, were they similar in terms of level of education, clinical or research experience, or level of responsibility in the piece of research being appraised? When there was more than one observer or collector, was there comparison of results from across the observers? Was the condition measured in the same way for all participants? | ☐ Yes<br>☐ No<br>☐ Unclear<br>☐ Not applicable |
| **8. Was there appropriate statistical analysis?**<br>Importantly, the numerator and denominator should be clearly reported, and percentages should be given with confidence intervals. The methods section should be detailed enough for reviewers to identify the analytical technique used and how specific variables were measured. Additionally, it is also important to assess the | ☐ Yes<br>☐ No<br>☐ Unclear<br>☐ Not applicable |
| appropriateness of the analytical strategy in terms of the assumptions associated with the approach as differing methods of analysis are based on differing assumptions about the data and how it will respond. |  |
| **9. Was the response rate adequate, and if not, was the low response rate managed appropriately?**<br>A large number of dropouts, refusals or “not founds” amongst selected subjects may diminish a study’s validity, as can a low response rates for survey studies. The authors should clearly discuss the response rate and any reasons for non-response and compare persons in the study to those not in the study, particularly with regards to their socio-demographic characteristics. If reasons for non-response appear to be unrelated to the outcome measured and the characteristics of non-responders are comparable to those who do respond in the study (addressed in question 5, coverage bias), the researchers may be able to justify a more modest response rate. | ☐ Yes<br>☐ No<br>☐ Unclear<br>☐ Not applicable |
| **Overall appraisal** | ☐ Low quality<br>☐ Medium quality<br>☐ High quality<br>☐ Seek further info |
| **Comments** | [Open text field] |

### Supplement 4 – Quality Appraisal

| S/No | Study Title | First author last name | Year of publication | Type of Study | Overall appraisal |
| --- | --- | --- | --- | --- | --- |
| 1 | Health care seeking behavior and patient delay in tuberculosis diagnosis | Almeida | 2015 | Analytical Cross-Sectional | High |
| 2 | The road to tuberculosis treatment in rural Nepal: A qualitative assessment of 26 journeys | Asbroek | 2008 | Qualitative | High |
| 3 | Delays to anti-tuberculosis treatment initiation among cases on directly observed treatment short course in districts of southwestern Ethiopia: a cross sectional study | Asres | 2019 | Analytical Cross-Sectional | High |
| 4 | Such a long journey: What health seeking pathways of patients with drug resistant tuberculosis in Mumbai tell us | Bhattacharya | 2019 | Qualitative | High |
| 5 | How do patients access the private sector in Chennai, India? An evaluation of delays in tuberculosis diagnosis | Bronner Murrison | 2016 | Cross-Sectional Study | High |
| 6 | Health-seeking behaviour and treatment delay in patients with pulmonary tuberculosis in Switzerland: some slip through the net | Christian | 2018 | Analytical Cross-Sectional | High |
| 7 | Delays in TB Diagnosis and Treatment Initiation in Burkina Faso during the COVID-19 Pandemic | Diallo | 2022 | Analytical Cross-Sectional | High |
| 8 | Health seeking behaviour and delayed management of tuberculosis patients in rural Bangladesh | Ehsanul Huq | 2018 | Analytical Cross-Sectional | High |
| 9 | Real-world colorectal cancer diagnostic pathways in Ontario, Canada: A | Guan | 2022 | Analytical Cross- | High |
|  | population-based study |  |  | Sectional |  |
| 10 | Patient pathways of tuberculosis care-seeking and treatment: an individual-level analysis of National Health Insurance data in Taiwan | Ku | 2020 | Cohort | High |
| 11 | Health-seeking behaviour for febrile illness in malaria-endemic Kolasib district, Mizoram, India | Lalchhuanawma | 2012 | Analytical Cross-Sectional | High |
| 12 | Patient pathways and delays to diagnosis and treatment of tuberculosis in an urban setting in Indonesia | Lestari | 2020 | Analytical Cross-Sectional | High |
| 13 | Health-seeking pathway and factors leading to delays in tuberculosis diagnosis in West Pokot County, Kenya: a grounded theory study | Mbuthia | 2018 | Qualitative | High |
| 14 | Delayed consultation among pulmonary tuberculosis patients: a cross sectional study of 10 DOTS districts of Ethiopia | Mesfin | 2009 | Analytical Cross-Sectional | High |
| 15 | Patterns and predictors of malaria care-seeking, diagnostic testing, and artemisinin-based combination therapy for children under five with fever in Northern Nigeria: a cross-sectional study | Millar | 2014 | Analytical Cross-Sectional | High |
| 16 | Demand for malaria rapid diagnostic test, health care-seeking behaviour, and drug use among rural community members with fever or malaria-like illness in Ebonyi state, Nigeria: a cross-sectional household survey | Omale | 2021 | Analytical Cross-Sectional | High |
| 17 | Care-seeking during fatal childhood illness in rural South Africa: a qualitative study | Price | 2021 | Qualitative | High |
| 18 | Effect of public-private interface agency in | Shah | 2020 | Analytical | High |
|  | Patna and Mumbai, India: Does it alter durations and delays in care seeking for drug-sensitive pulmonary tuberculosis? |  |  | Cross-Sectional |  |
| 19 | Healthcare-seeking behavior, treatment delays and its determinants among pulmonary tuberculosis patients in rural Nigeria: a cross-sectional study | Ukwaja | 2013 | Analytical Cross-Sectional | High |
| 20 | Tuberculosis patients' knowledge and beliefs about tuberculosis: a mixed methods study from the Pacific Island nation of Vanuatu | Viney | 2014 | Mixed Methods | High |
| 21 | Comparing patient care seeking pathways in three models of hospital and TB programme collaboration in China | Wei | 2013 | Analytical Cross-Sectional | High |
| 22 | Transaction costs of access to health care: Implications of the care-seeking pathways of tuberculosis patients for health system governance in Nigeria | Abimbola | 2015 | Cross-Sectional Study | Medium |
| 23 | Diagnostic delay amongst tuberculosis patients in Jogjakarta Province, Indonesia is related to the quality of services in DOTS facilities | Ahmad | 2011 | Cross-Sectional Study | Medium |
| 24 | Knowledge on the transmission, prevention and treatment of malaria among two endemic populations of Bangladesh and their health-seeking behaviour | Ahmed | 2009 | Cross-Sectional Study | Medium |
| 25 | Duration of intervals in the care seeking pathway for lung cancer in Bangladesh: A journey from symptoms triggering consultation to receipt of treatment | Ansar | 2021 | Cross-Sectional Study | Medium |
| 26 | Patient's site of first access to health system influences length of delay for tuberculosis treatment in Tajikistan | Aye | 2010 | Cross-Sectional Study | Medium |
| 27 | Care seeking and attitudes towards treatment compliance by newly enrolled tuberculosis patients in the district treatment programme in rural western Kenya: a qualitative study | Ayisi | 2011 | Qualitative Research | Medium |
| 28 | Men with sexually transmitted diseases in Bangkok: where do they go for treatment and why? | Benjarattanaporn | 1997 | Cross-Sectional Study | Medium |
| 29 | Pre-treatment delay and out of pocket expenses by notified new tuberculosis patients in an Indian mega city | Chadha | 2022 | Cross-Sectional Study | Medium |
| 30 | Care Seeking Behavior of Chest Symptomatics: A Community Based Study Done in South India after the Implementation of the RNTCP | Charles | 2010 | Cross-Sectional Study | Medium |
| 31 | Cost of seeking care for tuberculosis since the implementation of universal health coverage in Indonesia | Fuady | 2020 | Cross-Sectional Study | Medium |
| 32 | Care seeking behaviour and various delays in tuberculosis patients registered under RNTCP in Pune city | Gothankar | 2016 | Cross-Sectional Study | Medium |
| 33 | The relationship between poverty and healthcare seeking among patients hospitalized with acute febrile illnesses in Chittagong, Bangladesh | Herdman | 2016 | Cross-Sectional Study | Medium |
| 34 | Healthcare Seeking and Access to Care for Pneumonia, Sepsis, Meningitis, and Malaria in Rural Gambia | Hossain | 2022 | Cross-Sectional Study | Medium |
| 35 | Barriers on the pathway to survival for | Kallander | 2019 | Cross- | Medium |
|  | children dying from treatable illnesses in Inhambane province, Mozambique |  |  | Sectional Study |  |
| 36 | Study of factors leading to treatment delay in new sputum positive pulmonary tuberculosis patients and its impact on sputum conversion | Kumawat | 2022 | Cohort Study | Medium |
| 37 | Patients are paying too much for tuberculosis: a direct cost-burden evaluation in Burkina Faso | Laokri | 2013 | Cross-Sectional Study | Medium |
| 38 | Case-finding tuberculosis patients: diagnostic and treatment delays and their determinants | Maamari | 2008 | Cross-Sectional Study | Medium |
| 39 | How much do delayed healthcare seeking, delayed care provision, and diversion from primary care contribute to the transmission of STIs? | Mercer | 2007 | Cross-Sectional Study | Medium |
| 40 | Pulmonary tuberculosis in Patna, India: Durations, delays, and health care seeking behaviour among patients identified through household surveys | Mistry | 2017 | Cross-Sectional Study | Medium |
| 41 | Durations and Delays in Care Seeking, Diagnosis and Treatment Initiation in Uncomplicated Pulmonary Tuberculosis Patients in Mumbai, India | Mistry | 2016 | Mixed Methods | Medium |
| 42 | Patient navigation pathway and barriers to treatment seeking in cancer in India: A qualitative inquiry | Pati | 2013 | Mixed Methods | Medium |
| 43 | Care-seeking pathways, care challenges, and coping experiences of rural women living with rheumatoid arthritis in Odisha, India | Pati | 2019 | Qualitative Research | Medium |
| 44 | Factors associated with patient and health | Rajeswari | 2002 | Cross- | Medium |
|  | system delays in the diagnosis of tuberculosis in South India |  |  | Sectional Study |  |
| 45 | Patient treatment pathways of multidrug-resistant tuberculosis cases in coastal South India: Road to a drug resistant tuberculosis center | Rathi | 2021 | Cross-Sectional Study | Medium |
| 46 | Care seeking behaviour and diagnostic processes in patients with smear-positive pulmonary tuberculosis in Malawi | Salaniponi | 2000 | Cross-Sectional Study | Medium |
| 47 | A comparison of patient treatment pathways among multidrug-resistant and drug-sensitive TB cases in Delhi, India: A cross-sectional study | Sharma | 2020 | Cross-Sectional Study | Medium |
| 48 | From fever to anti-malarial: the treatment-seeking process in rural Senegal | Smith | 2010 | Cross-Sectional Study | Medium |
| 49 | Pathways to the first contact with specialist mental health care | Steel | 2006 | Cross-Sectional Study | Medium |
| 50 | Health care seeking patterns of rifampicin-resistant tuberculosis patients in Harare, Zimbabwe: A prospective cohort study | Tadokera | 2021 | Mixed Methods | Medium |
| 51 | Understanding health care-seeking behaviour of the tribal population in India among those with presumptive TB symptoms | Thomas | 2021 | Cross-Sectional Study | Medium |
| 52 | Diagnostic pathways and direct medical costs incurred by new adult pulmonary tuberculosis patients prior to anti-tuberculosis treatment “ Tamil Nadu, India | Veesa | 2018 | Cross-Sectional Study | Medium |
| 53 | Analysis of care-seeking pathways of tuberculosis patients in Guangxi, China, with and without decentralised tuberculosis services | Wei | 2009 | Cross-Sectional Study | Medium |
| 54 | Patient care pathways under the model of integrating tuberculosis service with general hospitals in China | Wei | 2013 | Cross-Sectional Study | Medium |
| 55 | The pathway to diagnosis and treatment for surgically managed lung cancer patients | White | 2020 | Cross-Sectional Study | Medium |
| 56 | Healthcare utilization for common infectious disease syndromes in Soweto and Klerksdorp, South Africa | Wong | 2018 | Cross-Sectional Study | Medium |
| 57 | Patients pathways to tuberculosis diagnosis and treatment in a fragmented health system: a qualitative study from a south Indian district | Yellappa | 2017 | Qualitative Research | Medium |
| 58 | Patient pathway analysis of tuberculosis diagnostic delay: a multicentre retrospective cohort study in China | Zhang | 2021 | Cohort Study | Medium |
| 59 | Care Seeking Behavior of the TB Patients who were Registered in an Urban Government Tuberculosis Control in Chennai, Tamilnadu, India | Ananthakrishnan | 2012 | Cross-Sectional Study | Low |
| 60 | Tuberculosis pathways to care and transmission of multidrug resistance in India | Atre | 2022 | Mixed Methods | Low |
| 61 | Health seeking behaviour, health system experience and tuberculosis case finding in Gambians with cough | Kasse | 2006 | Cross-Sectional Study | Low |
| 62 | Idiopathic pulmonary fibrosis: Physician and patient perspectives on the pathway to care from symptom recognition to diagnosis and disease burden | Lancaster | 2021 | Cross-Sectional Study | Low |
| 63 | Pathways and costs of care for patients | Shete | 2015 | Cross- | Low |
|  | with tuberculosis symptoms in rural Uganda |  |  | Sectional Study |  |
| 64 | Health seeking and knowledge about tuberculosis among persons with pulmonary symptoms and tuberculosis cases in Bangalore slums | Suganthi | 2008 | Cross-Sectional Study | Low |
| 65 | Health-seeking behavior of COVID-19 cases during the first eight weeks of the outbreak in Singapore: differences between local community and imported cases and having visits to single or multiple healthcare providers | Tay | 2022 | Cross-Sectional Study | Low |
| 66 | A Cross-Sectional Descriptive Study to Ascertain Factors Influencing Delay in Diagnosis among Newly Diagnosed Pulmonary Tuberculosis Patients | Thomas | 2022 | Cross-Sectional Study | Low |

## References

1. World Health Organization. Tracking universal health coverage: 2023 global monitoring report: World Health Organization; 2023.

2. World Health Organization. Global tuberculosis report 2023. 2023.

3. Joint United Nations Programme on HIV/AIDS. The path that ends AIDS: 2023 UNAIDS global AIDS update. 2023.

4. Faust L, Naidoo P, Caceres-Cardenas G, et al. Improving measurement of tuberculosis care cascades to enhance people-centred care. The Lancet Infectious Diseases 2023.

5. MacPherson P, Houben RM, Glynn JR, Corbett EL, Kranzer K. Pre-treatment loss to follow-up in tuberculosis patients in low-and lower-middle-income countries and high-burden countries: a systematic review and meta-analysis. Bulletin of the World Health Organization 2013; 92: 126–38.

6. Arokiasamy P, Uttamacharya, Kowal P, et al. Chronic noncommunicable diseases in 6 low-and middle-income countries: findings from wave 1 of the World Health Organization’s study on global Ageing and adult health (SAGE). American journal of epidemiology 2017; 185(6): 414–28.

7. Johnson CC, Fonner V, Sands A, et al. To err is human, to correct is public health: a systematic review examining poor quality testing and misdiagnosis of HIV status. Journal of the International AIDS Society 2017; 20: 21755.

8. Mwenesi H, Harpham T, Snow RW. Child malaria treatment practices among mothers in Kenya. Social science & medicine 1995; 40(9): 1271–7.

9. Räisänen U, Hunt K. The role of gendered constructions of eating disorders in delayed help-seeking in men: a qualitative interview study. BMJ open 2014; 4(4): e004342.

10. Storla DG, Yimer S, Bjune GA. A systematic review of delay in the diagnosis and treatment of tuberculosis. BMC public health 2008; 8: 1–9.

11. Oga-Omenka C, Tseja-Akinrin A, Sen P, et al. Factors influencing diagnosis and treatment initiation for multidrug-resistant/rifampicin-resistant tuberculosis in six sub-Saharan African countries: a mixed-methods systematic review. BMJ global health 2020; 5(7): e002280.

12. Mavhu W, Dauya E, Bandason T, et al. Chronic cough and its association with TB–HIV co-infection: factors affecting help-seeking behaviour in Harare, Zimbabwe. Tropical Medicine & International Health 2010; 15(5): 574–9.

13. Allen H, Wright BJ, Harding K, Broffman L. The role of stigma in access to health care for the poor. The Milbank Quarterly 2014; 92(2): 289–318.

14. Cai J, Wang X, Ma A, Wang Q, Han X, Li Y. Factors associated with patient and provider delays for tuberculosis diagnosis and treatment in Asia: a systematic review and meta-analysis. PloS one 2015; 10(3): e0120088.

15. Stallworthy G, Dias HM, Pai M. Quality of tuberculosis care in the private health sector. Journal of clinical tuberculosis and other mycobacterial diseases 2020; 20: 100171.

16. Carrillo JE, Carrillo VA, Perez HR, Salas-Lopez D, Natale-Pereira A, Byron AT. Defining and targeting health care access barriers. Journal of health care for the poor and underserved 2011; 22(2): 562–75.

17. Tanimura T, Jaramillo E, Weil D, Raviglione M, Lönnroth K. Financial burden for tuberculosis patients in low-and middle-income countries: a systematic review. European Respiratory Journal 2014; 43(6): 1763–75.

18. Organization WH. The END TB strategy: World Health Organization; 2015.

19. World Health Organization. Interim report: placing people and communities at the centre of health services: WHO global strategy on integrated people-centred health services 2016-2026: executive summary: World Health Organization, 2015.

20. World Health Organization. World Health Assembly adopts framework on integrated people-centred health services. World Health Organization 2016.

21. Odone A, Roberts B, Dara M, Van Den Boom M, Kluge H, McKee M. People-and patient-centred care for tuberculosis: models of care for tuberculosis. The international journal of tuberculosis and lung disease 2018; 22(2): 133–8.

22. Shaller D. Patient-centered care: what does it take?: Commonwealth Fund New York; 2007.

23. World Health Organization. Compendium of data and evidence-related tools for use in TB planning and programming. Compendium of data and evidence-related tools for use in TB planning and programming; 2021.

24. Oliver A, Mossialos E. Equity of access to health care: outlining the foundations for action. Journal of Epidemiology & Community Health 2004; 58(8): 655–8.

25. Oga-Omenka C, Bada F, Agbaje A, Dakum P, Menzies D, Zarowsky C. Ease and equity of access to free DR-TB services in Nigeria-a qualitative analysis of policies, structures and processes. International Journal for Equity in Health 2020; 19: 1–13.

26. Daniels N. Equity of access to health care: some conceptual and ethical issues. The Milbank Memorial Fund Quarterly Health and Society 1982: 51–81.

27. Huria L, Lestari BW, Saptiningrum E, et al. Care pathways of individuals with tuberculosis before and during the COVID-19 pandemic in Bandung, Indonesia. PLOS Global Public Health 2024; 4(1): e0002251.

28. Lestari BW, McAllister S, Hadisoemarto PF, et al. Patient pathways and delays to diagnosis and treatment of tuberculosis in an urban setting in Indonesia. Lancet Regional Health-Western Pacific 2020; 5.

29. Oga-Omenka C, Rosapep L, Baruwa E, et al. Individual journeys to tuberculosis care in Nigeria’s private sector during the COVID-19 pandemic. BMJ Global Health 2024; 9(1): e013124.

30. Pinder R, Petchey R, Shaw S, Carter Y. What’s in a care pathway? Towards a cultural cartography of the new NHS. Sociology of health & illness 2005; 27(6): 759–79.

31. Lismont J, Janssens A-S, Odnoletkova I, vanden Broucke S, Caron F, Vanthienen J. A guide for the application of analytics on healthcare processes: A dynamic view on patient pathways. Computers in biology and medicine 2016; 77: 125–34.

32. Richter P, Schlieter H. Understanding patient pathways in the context of integrated health care services-implications from a scoping review. 2019.

33. Gartner J-B, Abasse KS, Bergeron F, Landa P, Lemaire C, Côté A. Definition and conceptualization of the patient-centered care pathway, a proposed integrative framework for consensus: a concept analysis and systematic review. BMC health services research 2022; 22(1): 558.

34. Hanson CL, Osberg M, Brown J, Durham G, Chin DP. Conducting patient-pathway analysis to inform programming of tuberculosis services: methods. The Journal of infectious diseases 2017; 216(suppl_7): S679–S85.

35. Hanson C, Osberg M, Brown J, Durham G, Chin DP. Finding the missing patients with tuberculosis: lessons learned from patient-pathway analyses in 5 countries. The Journal of infectious diseases 2017; 216(suppl_7): S686–S95.

36. Li Y, Ehiri J, Tang S, et al. Factors associated with patient, and diagnostic delays in Chinese TB patients: a systematic review and meta-analysis. BMC medicine 2013; 11: 1–15.

37. Obsa MS, Daga WB, Wosene NG, et al. Treatment seeking delay and associated factors among tuberculosis patients attending health facility in Ethiopia from 2000 to 2020: a systematic review and meta analysis. PloS one 2021; 16(7): e0253746.

38. Ukwaja KN, Alobu I, Nweke CO, Onyenwe EC. Healthcare-seeking behavior, treatment delays and its determinants among pulmonary tuberculosis patients in rural Nigeria: a cross-sectional study. BMC health services research 2013; 13: 1–9.

39. Almeida CPBd, Skupien EC, Silva DR. Health care seeking behavior and patient delay in tuberculosis diagnosis. Cadernos de saude publica 2015; 31(2): 321–30.

40. Fuady A, Houweling TAJ, Mansyur M, Burhan E, Richardus JH. Cost of seeking care for tuberculosis since the implementation of universal health coverage in Indonesia. BMC Health Services Research 2020; 20(502).

41. Camfield L, Streuli N, Woodhead M. Children’s well-being in developing countries: A conceptual and methodological review. The European Journal of Development Research 2010; 22: 398–416.

42. Wang J, Lloyd-Evans B, Giacco D, et al. Social isolation in mental health: a conceptual and methodological review. Social psychiatry and psychiatric epidemiology 2017; 52: 1451–61.

43. Kallio H, Pietilä AM, Johnson M, Kangasniemi M. Systematic methodological review: developing a framework for a qualitative semi-structured interview guide. Journal of advanced nursing 2016; 72(12): 2954–65.

44. Frederiksen L, Phelps SF, Kimmons R. Introduction to literature reviews. Rapid academic writing [document on the Internet] c2018 [cited 2021 Oct 13] Available from: https://edtechbooks.org/pdfs/mobile/rapidwriting/lit_rev_intro.pdf 2018.

45. Hulland J. Conceptual review papers: revisiting existing research to develop and refine theory. AMS Review 2020; 10(1): 27–35.

46. Jit M, Ng DHL, Luangasanatip N, et al. Quantifying the economic cost of antibiotic resistance and the impact of related interventions: rapid methodological review, conceptual framework and recommendations for future studies. BMC medicine 2020; 18: 1–14.

47. Campbell M, Egan M, Lorenc T, et al. Considering methodological options for reviews of theory: illustrated by a review of theories linking income and health. Systematic reviews 2014; 3: 1–11.

48. Mbuagbaw L, Lawson DO, Puljak L, Allison DB, Thabane L. A tutorial on methodological studies: the what, when, how and why. BMC Medical Research Methodology 2020; 20: 1–12.

49. Munn Z, Stern C, Aromataris E, Lockwood C, Jordan Z. What kind of systematic review should I conduct? A proposed typology and guidance for systematic reviewers in the medical and health sciences. BMC medical research methodology 2018; 18: 1–9.

50. Lilford RJ, Richardson A, Stevens A, et al. Issues in methodological research: perspectives from researchers and commissioners. 2001.

51. Joanna Briggs Institute. Critical Appraisal Tools. 2020. https://jbi.global/critical-appraisal-tools.

52. Hong QN, Fàbregues S, Bartlett G, et al. The Mixed Methods Appraisal Tool (MMAT) version 2018 for information professionals and researchers. Education for information 2018; 34(4): 285–91.

53. Yang W-T, Gounder CR, Akande T, et al. Barriers and delays in tuberculosis diagnosis and treatment services: does gender matter? Tuberculosis research and treatment 2014; 2014.

54. Subbaraman R, Jhaveri T, Nathavitharana RR. Closing gaps in the tuberculosis care cascade: an action-oriented research agenda. Journal of clinical tuberculosis and other mycobacterial diseases 2020; 19: 100144.

55. Mugavero MJ, Amico KR, Horn T, Thompson MA. The state of engagement in HIV care in the United States: from cascade to continuum to control. Clinical infectious diseases 2013; 57(8): 1164–71.

56. Oga-Omenka C, Boffa J, Kuye J, Dakum P, Menzies D, Zarowsky C. Understanding the gaps in DR-TB care cascade in Nigeria: A sequential mixed-method study. Journal of Clinical Tuberculosis and Other Mycobacterial Diseases 2020; 21: 100193.

57. Baru RV. Disease and Suffering: Towards a Framework for Understanding Health Seeking Behavior. Indian Anthropologist 2005: 45–52.

58. Lestari BW, McAllister S, Hadisoemarto PF, et al. Patient pathways and delays to diagnosis and treatment of tuberculosis in an urban setting in Indonesia. The Lancet Regional Health–Western Pacific 2020; 5.

59. Ahmad RA, Mahendradhata Y, Utarini A, de Vlas SJ. Diagnostic delay amongst tuberculosis patients in Jogjakarta Province, Indonesia is related to the quality of services in DOTS facilities. Tropical Medicine & International Health 2011; 16(4): 412–23.

60. Ansar A, Lewis V, McDonald CF, Liu C, Rahman MA. Duration of intervals in the care seeking pathway for lung cancer in Bangladesh: A journey from symptoms triggering consultation to receipt of treatment. PloS one 2021; 16(9): e0257301.

61. Asres A, Jerene D, Deressa W. Delays to anti-tuberculosis treatment intiation among cases on directly observed treatment short course in districts of southwestern Ethiopia: a cross sectional study. BMC Infect Dis 2019; 19(1): 481.

62. Atre SR, Jagtap JD, Faqih MI, et al. Tuberculosis pathways to care and transmission of multidrug resistance in India. American Journal of Respiratory and Critical Care Medicine 2022; 205(2): 233–41.

63. Ayisi JG, Hoog AHv, Agaya JA, et al. Care seeking and attitudes towards treatment compliance by newly enrolled tuberculosis patients in the district treatment programme in rural western Kenya: a qualitative study. BMC Public Health 2011; 11(515).

64. Bronner Murrison L, Ananthakrishnan R, Swaminathan A, et al. How do patients access the private sector in Chennai, India? An evaluation of delays in tuberculosis diagnosis. Int J Tuberc Lung Dis 2016; 20(4): 544–51.

65. Diallo A, Combary A, Veronese V, et al. Delays in TB Diagnosis and Treatment Initiation in Burkina Faso during the COVID-19 Pandemic. Trop Med Infect Dis 2022; 7(9).

66. Gothankar JS, Patil UP, Gaikwad SR, Kamble SB. Care seeking behaviour and various delays in tuberculosis patients registered under RNTCP in Pune city. Indian Journal of Community Health 2016; 28(1): 34–9.

67. Guan Z, Webber C, Flemming JA, et al. Real-world colorectal cancer diagnostic pathways in Ontario, Canada: A population-based study. European Journal of Cancer Care 2022; 31(5): 1-13.

68. Herdman MT, Maude RJ, Chowdhury MS, et al. The relationship between poverty and healthcare seeking among patients hospitalized with acute febrile illnesses in Chittagong, Bangladesh. PLoS One 2016; 11(4): e0152965.

69. Kasse Y, Jasseh M, Corrah T, et al. Health seeking behaviour, health system experience and tuberculosis case finding in Gambians with cough. BMC Public Health 2006; 6(143).

70. Kumawat A, Chakraborti A, Kumar S, et al. Study of factors leading to treatment delay in new sputum positive pulmonary tuberculosis patients and its impact on sputum conversion. Tropical Doctor.

71. Lancaster L, Bonella F, Inoue Y, et al. Idiopathic pulmonary fibrosis: Physician and patient perspectives on the pathway to care from symptom recognition to diagnosis and disease burden. Respirology (Carlton, Vic) 2022; 27(1): 66–75.

72. Laokri S, Drabo MK, Weil O, Kafando B, Dembélé SM, Dujardin B. Patients are paying too much for tuberculosis: a direct cost-burden evaluation in Burkina Faso. PLoS One 2013; 8(2): e56752.

73. Maamari F. Case-finding tuberculosis patients: diagnostic and treatment delays and their determinants. Eastern Mediterranean Health Journal 2008; 14(3): 531–45.

74. Mercer CH, Sutcliffe L, Johnson AM, et al. How much do delayed healthcare seeking, delayed care provision, and diversion from primary care contribute to the transmission of STIs? SEXUALLY TRANSMITTED INFECTIONS 2007; 83(5): 400–5.

75. Mistry N, Lobo E, Shah S, Rangan S, Dholakia Y. Pulmonary tuberculosis in Patna, India: Durations, delays, and health care seeking behaviour among patients identified through household surveys. Journal of Epidemiology and Global Health 2017; 7(4): 241-8.

76. Mistry N, Rangan S, Dholakia Y, Lobo E, Shah S, Patil A. Durations and Delays in Care Seeking, Diagnosis and Treatment Initiation in Uncomplicated Pulmonary Tuberculosis Patients in Mumbai, India. PLoS One 2016; 11(3).

77. Priya R, Kalpita S, Bhaskaran U, Abhinav P, Abhirami N. Patient treatment pathways of multidrug-resistant tuberculosis cases in coastal South India: road to a drug resistant tuberculosis center [version 5; peer review: 2 approved, 1 approved with reservations]. F1000Research 2021; 8(498).

78. Rajeswari R, Chandrasekaran V, Suhadev M, Sivasubramaniam S, Sudha G, Renu G. Factors associated with patient and health system delays in the diagnosis of tuberculosis in South India. International Journal of Tuberculosis and Lung Disease 2002; 6(9): 789–95.

79. Salaniponi FM, Harries AD, Banda HT, et al. Care seeking behaviour and diagnostic processes in patients with smear-positive pulmonary tuberculosis in Malawi. Int J Tuberc Lung Dis 2000; 4(4): 327–32.

80. Shah S, Shah S, Rangan S, et al. Effect of public-private interface agency in Patna and Mumbai, India: Does it alter durations and delays in care seeking for drug-sensitive pulmonary tuberculosis? Gates Open Res 2020; 4: 32.

81. Sharma N, Bhatnagar A, Basu S, et al. A comparison of patient treatment pathways among multidrug-resistant and drug-sensitive TB cases in Delhi, India: A cross-sectional study. Indian J Tuberc 2020; 67(4): 502-8.

82. Steel Z, McDonald R, Silove D, et al. Pathways to the first contact with specialist mental health care. Australian & New Zealand Journal of Psychiatry 2006; 40(4): 347–54.

83. Tay M, Ang L, Wei W, Lee VJM, Leo Y, Toh MPHS. Health-seeking behavior of COVID- 19 cases during the first eight weeks of the outbreak in Singapore: differences between local community and imported cases and having visits to single or multiple healthcare providers. BMC Public Health 2022; 22(239).

84. Thomas N, Rajalingam R, Vallabhaneni V, Varghese J. A Cross-Sectional Descriptive Study to Ascertain Factors Influencing Delay in Diagnosis among Newly Diagnosed Pulmonary Tuberculosis Patients. Indian Journal of Respiratory Care 2022; 11(4): 341–8.

85. Veesa KS, John KR, Moonan PK, et al. Diagnostic pathways and direct medical costs incurred by new adult pulmonary tuberculosis patients prior to anti-tuberculosis treatment - Tamil Nadu, India. PLoS One 2018; 13(2): e0191591.

86. Viney KA, Johnson P, Tagaro M, et al. Tuberculosis patients’ knowledge and beliefs about tuberculosis: a mixed methods study from the Pacific Island nation of Vanuatu. BMC Public Health 2014; 14.

87. Wei X, Zou G, Yin J, Walley J, Sun Q. Comparing patient care seeking pathways in three models of hospital and TB programme collaboration in China. BMC Infectious Diseases 2013; 13(93).

88. Wei XL, Liang XY, Walley JD, Liu FY, Dong BQ. Analysis of care-seeking pathways of tuberculosis patients in Guangxi, China, with and without decentralised tuberculosis services. International Journal of Tuberculosis and Lung Disease 2009; 13(4): 514–20.

89. White V, Bergin RJ, Thomas RJ, Whitfield K, Weller D. The pathway to diagnosis and treatment for surgically managed lung cancer patients. Family practice 2020; 37(2): 234–41.

90. Yellappa V, Lefèvre P, Battaglioli T, Devadasan N, Van der Stuyft P. Patients pathways to tuberculosis diagnosis and treatment in a fragmented health system: a qualitative study from a south Indian district. BMC public health 2017; 17(1): 1–10.

91. Zhang L, Weng T, Wang H, et al. Patient pathway analysis of tuberculosis diagnostic delay: a multicentre retrospective cohort study in China. Clinical Microbiology and Infection 2021; 27(7): 1000–6.

92. Abimbola S, Ukwaja KN, Onyedum CC, Negin J, Jan S, Martiniuk AL. Transaction costs of access to health care: Implications of the care-seeking pathways of tuberculosis patients for health system governance in Nigeria. Glob Public Health 2015; 10(9): 1060–77.

93. Asbroek AHAt, Bijlsma MW, Malla P, Shrestha B, Delnoij DMJ. The road to tuberculosis treatment in rural Nepal: a qualitative assessment of 26 journeys. BMC Health Services Research 2008; 8(7).

94. Aye R, Wyss K, Abdualimova H, Saidaliev S. Patient’s site of first access to health system influences length of delay for tuberculosis treatment in Tajikistan. BMC Health Services Research 2010; 10(10).

95. Benjarattanaporn P, Lindan CP, Mills S, et al. Men with sexually transmitted diseases in Bangkok: where do they go for treatment and why? Aids 1997; 11(Supplement1): S87–S95.

96. Bhattacharya Chakravarty A, Rangan S, Dholakia Y, et al. Such a long journey: What health seeking pathways of patients with drug resistant tuberculosis in Mumbai tell us. PLoS One 2019; 14(1): e0209924.

97. Kallander K, Counihan H, Cerveau T, Mbofana F. Barriers on the pathway to survival for children dying from treatable illnesses in Inhambane province, Mozambique. Journal of Global Health 2019; 9(1).

98. Mesfin MM, Newell JN, Walley JD, Gessessew A, Madeley RJ. Delayed consultation among pulmonary tuberculosis patients: a cross sectional study of 10 DOTS districts of Ethiopia. BMC Public Health 2009; 9(53).

99. Millar KR, McCutcheon J, Coakley EH, et al. Patterns and predictors of malaria care-seeking, diagnostic testing, and artemisinin-based combination therapy for children under five with fever in Northern Nigeria: a cross-sectional study. Malaria Journal 2014; 13(447).

100. Pati S, Sahoo KC, Samal M, et al. Care-seeking pathways, care challenges, and coping experiences of rural women living with rheumatoid arthritis in Odisha, India. Primary Health Care Research & Development 2019; 20.

101. Renthlei L, Murhekar MV. Health-seeking behaviour for febrile illness in malaria-endemic Kolasib district, Mizoram, India. International Health (RSTMH) 2012; 4(4): 314–9.

102. Shete PB, Haguma P, Miller CR, et al. Pathways and costs of care for patients with tuberculosis symptoms in rural Uganda. International Journal of Tuberculosis and Lung Disease 2015; 19(8): 912–7.

103. Tadokera R, Huo S, Theron G, Timire C, Manyau-Makumbirofa S, Metcalfe JZ. Health care seeking patterns of rifampicin-resistant tuberculosis patients in Harare, Zimbabwe: A prospective cohort study. PLoS One 2021; 16(7).

104. Thomas BE, Thiruvengadam K, Raghavi S, et al. Understanding health care-seeking behaviour of the tribal population in India among those with presumptive TB symptoms. PLoS One 2021; 16(5).

105. Wei X, Yin J, Zou G, et al. Patient care pathways under the model of integrating tuberculosis service with general hospitals in China. Tropical Medicine & International Health 2013; 18(11): 1392–9.

106. Wong K, Mollendorf Cv, Martinson N, et al. Healthcare utilization for common infectious disease syndromes in Soweto and Klerksdorp, South Africa. Pan African Medical Journal 2018; 30: 271.

107. Almeida CP, Skupien EC, Silva DR. Health care seeking behavior and patient delay in tuberculosis diagnosis. Cad Saude Publica 2015; 31(2): 321–30.

108. Ananthakrishnan R, Jeyaraju AR, Palani G, Sathiyasekaran BWC. Care Seeking Behavior of the TB Patients who were Registered in an Urban Government Tuberculosis Control in Chennai, Tamilnadu, India. Journal of Clinical and Diagnostic Research 2012; 6(6): 990–3.

109. Charles N, Thomas B, Watson B, Raja Sakthivel M, Chandrasekeran V, Wares F. Care seeking behavior of chest symptomatics: a community based study done in South India after the implementation of the RNTCP. PLoS One 2010; 5(9).

110. Suganthi P, Chadha VK, Ahmed J, et al. Health seeking and knowledge about tuberculosis among persons with pulmonary symptoms and tuberculosis cases in Bangalore slums. International Journal of Tuberculosis and Lung Disease 2008; 12(11): 1268–73.

111. Smith LA, Bruce J, Gueye L, et al. From fever to anti-malarial: the treatment-seeking process in rural Senegal. Malaria Journal 2010; 9(333).

112. Ehsanul Huq K, Moriyama M, Zaman K, et al. Health seeking behaviour and delayed management of tuberculosis patients in rural Bangladesh. BMC Infect Dis 2018; 18(1): 515.

113. Ahmed SM, Rashidul H, Ubydul H, Awlad H. Knowledge on the transmission, prevention and treatment of malaria among two endemic populations of Bangladesh and their health-seeking behaviour. Malaria Journal 2009; 8(173).

114. Auer C, Kiefer S, Zuske M, et al. Health-seeking behaviour and treatment delay in patients with pulmonary tuberculosis in Switzerland: some slip through the net. Swiss Med Wkly 2018; 148: w14659.

115. Chadha VK, Praseeja P, Srivastava R, et al. Pre-treatment delay and out of pocket expenses by notified new tuberculosis patients in an Indian mega city. Indian J Tuberc 2022; 69(4): 446–52.

116. Ku CC, Chen CC, Dixon S, Lin HH, Dodd PJ. Patient pathways of tuberculosis care-seeking and treatment: an individual-level analysis of National Health Insurance data in Taiwan. BMJ Global Health 2020; 5(6).

117. Ukwaja KN, Alobu I, Nweke CO, Onyenwe EC. Healthcare-seeking behavior, treatment delays and its determinants among pulmonary tuberculosis patients in rural Nigeria: a cross-sectional study. BMC Health Services Research 2013; 13(25).

118. Pati S, Hussain MA, Chauhan AS, Mallick D, Nayak S. Patient navigation pathway and barriers to treatment seeking in cancer in India: a qualitative inquiry. Cancer Epidemiology 2013; 37(6): 973–8.

119. Hossain I, Hill P, Bottomley C, et al. Healthcare seeking and access to care for pneumonia, sepsis, meningitis, and malaria in rural Gambia. American Journal of Tropical Medicine and Hygiene 2022; 106(2): 446–53.

120. Omale UI, Oka OU, Okeke IM, et al. Demand for malaria rapid diagnostic test, health care-seeking behaviour, and drug use among rural community members with fever or malaria-like illness in Ebonyi state, Nigeria: a cross-sectional household survey. BMC Health Services Research 2021; 21(1).

121. Price J, Willcox M, Dlamini V, et al. Care-seeking during fatal childhood illness in rural South Africa: a qualitative study. BMJ Open 2021; 11(4).

